# Exploratory Symptom Profiles and Clinical Correlates in Adults with Attention-Deficit/Hyperactivity Disorder: A Retrospective Study Using the Wender-Reimherr Adult Attention Deficit Disorder Scale

**DOI:** 10.64898/2026.10.29.26366810

**Authors:** Taini Theuerkorn, Nicole Mauche, Jue Huang, Christian Makiol, Maria Strauß

## Abstract

**Background:** Emotional dysregulation (ED) is a clinically relevant feature of adult attention-deficit/hyperactivity disorder (ADHD), but its relationship to clinical burden remains incompletely characterized. We aimed to describe latent symptom profiles and compare clinical and sociodemographic characteristics of adults assigned to classes with elevated probabilities in emotion-related domains with those assigned to the remaining classes.

**Methods:** This retrospective cross-sectional study included 749 adults with ADHD from a specialized outpatient clinic. Latent class analysis was performed on 28 items of the clinician-rated Wender-Reimherr Adult Attention Deficit Disorder Scale (WRAADDS). Classes with elevated probabilities in temper, affective lability, and emotional overreactivity were grouped post hoc as an exploratory ADHD+ED group; the remaining classes formed the ADHD group. These labels do not denote clinical diagnoses. Sensitivity analyses examined alternative groupings and adjustment for core ADHD symptom severity.

**Results:** A six-class solution was retained. Under the primary grouping, 60.9% of participants were assigned to ED-related classes (24.6%–77.0% across alternative groupings). The ADHD+ED group showed greater psychological distress, depressive symptoms, and functional impairment; these differences persisted after adjustment for core symptom severity. Differences on several subscales of the Conners’ Adult ADHD Rating Scales (CAARS) also remained, whereas differences in broader ADHD self-report scores and the number of comorbid diagnoses did not. The small difference in drug-use screening scores was not robust in sensitivity analyses; alcohol-use screening scores and sociodemographic characteristics did not differ significantly.

**Conclusions:** Assignment to the exploratory ED-related group was associated with greater psychological distress, depressive symptoms, and functional impairment. The proportion assigned depended on the grouping and should not be interpreted as an ED prevalence estimate. The cross-sectional design limits causal inference; the single-center sample and exclusion of severe psychiatric comorbidities limit generalizability; and the shared WRAADDS basis limits independent interpretation of WRAADDS-derived group differences. Future research should use independent, dimensional measures of ED and longitudinal designs, and should evaluate interventions targeting emotion regulation in adults with ADHD.

## 1 Introduction

Attention-deficit/hyperactivity disorder (ADHD) is a childhood-onset neurodevelopmental disorder characterized by persistent patterns of inattention and/or hyperactivity-impulsivity (Falkai et al., 2025). In adulthood, prevalence estimates depend on how the disorder is defined. A global meta-analysis estimated the 2020 prevalence, adjusted for the global demographic structure, at 2.58% for persistent adult ADHD with childhood onset and 6.76% for symptomatic adult ADHD irrespective of childhood onset (Song et al., 2021).

Diagnosis of adult ADHD is based on a comprehensive clinical assessment of current and childhood symptoms, developmental history, and functional impairment. Informant information can provide additional support when available; rating scales may supplement but should not replace the clinical interview (Falkai et al., 2025; Adamou et al., 2024). Differences in diagnostic criteria and assessment procedures, as well as symptom overlap with other psychiatric disorders, can complicate differential diagnosis and comparisons across studies (Adamou et al., 2024; Choi et al., 2022). Symptom expression can change across adulthood: overt motor hyperactivity may become less prominent, while inattention and subjective restlessness can remain salient (Falkai et al., 2025; Cortese et al., 2025). Adults with ADHD have higher rates of substance use, mood, anxiety and related disorders (including somatoform disorders), and personality disorders than adults without ADHD (Fayyad et al., 2017; Choi et al., 2022). In a nationally representative Canadian sample, lifetime suicide attempts were reported more often by adults with ADHD than by adults without ADHD (14.0% vs. 2.7%; Fuller-Thomson et al., 2022).

The World Health Organization (WHO) classifies ADHD (F90.0) within the group of hyperkinetic disorders in the International Classification of Diseases, 10th Revision (ICD-10; Dilling and Freyberger, 2019). The Diagnostic and Statistical Manual of Mental Disorders, Fifth Edition, Text Revision (DSM-5-TR; Falkai et al., 2025), lists low frustration tolerance, irritability, and mood lability as associated features supporting diagnosis, although they are not diagnostic criteria; these features are commonly considered aspects of emotional dysregulation (ED; Faraone et al., 2019). In the ICD-11, the WHO also describes ED and low frustration tolerance as additional clinical features of ADHD, rather than diagnostic requirements (World Health Organization, 2024). Although ED is increasingly recognized as clinically relevant in adult ADHD, it lacks a standardized definition, and studies use overlapping but non-equivalent constructs and assessment measures, limiting direct comparisons across findings (Martz et al., 2023; Soler-Gutiérrez et al., 2023).

Clinic-based studies have reported impairing ED in 34% to 70% of adults with ADHD, although population-based studies are needed (Shaw et al., 2014). Shaw et al. (2014) describe ED as a dimensional trait that is not unique to ADHD, rather than as a categorical diagnosis. Their conceptualization includes emotional responses that are excessive relative to social norms or inappropriate to the context, rapid and poorly controlled shifts in emotion, and atypical allocation of attention to emotional stimuli. Related concepts in adult ADHD research include deficient emotional self-regulation and emotional impulsivity, which may manifest as mood lability, temper outbursts, and irritability (Barkley and Fischer, 2010; Surman et al., 2013; Faraone et al., 2019). In a controlled family study, deficient emotional self-regulation was more common among adults with ADHD than among controls and was associated with lower quality of life, poorer social adjustment, lower likelihood of being married, and higher risks of traffic accidents and arrests (Surman et al., 2013).

The Utah criteria represent an alternative conceptualization of adult ADHD that considers features beyond the traditional DSM symptom domains (Wender et al., 1981). Based on this framework, the Wender-Reimherr Adult Attention Deficit Disorder Scale (WRAADDS) assesses, in addition to attention difficulties, hyperactivity/restlessness, and impulsivity, the domains of disorganization and three emotion-related domains: temper, affective lability, and emotional overreactivity (Marchant et al., 2013). These emotion-related domains overlap with, but are not equivalent to, ED as a broader construct. The Conners’ Adult ADHD Rating Scales (CAARS) include an “Impulsivity/Emotional Lability” scale (Christiansen et al., 2014), while Barkley and Fischer (2010) found, in children with hyperactivity followed into adulthood, that emotional impulsiveness contributed to impairment in major life activities beyond ADHD symptom severity.

Person-centered studies have characterized heterogeneity in ADHD using different measures and classification methods. In a large sample of children with ADHD, latent class analysis of inattentive, hyperactive/impulsive, and ED symptoms identified five profiles, three of which included ED; these profiles were associated with more comorbidity, greater parent-rated everyday executive-function difficulties, and greater functional impairment. The findings were cross-sectional and based primarily on parent ratings (Yue et al., 2022). In adults, WRAADDS-based analyses have distinguished an inattentive presentation from an emotional dysregulation presentation (Reimherr et al., 2015). In a subsequent analysis of data from eight adult clinical trials, the original factor structure fit poorly, but an alternative two-factor solution again distinguished these presentations (Reimherr et al., 2020). Cluster- and factor-based studies in adults with ADHD have also described profiles characterized by ED or emotional instability (Hirsch et al., 2019; Pallucchini et al., 2021; Martz et al., 2023). A recent latent profile analysis based on emotion-regulation measures identified well-adapted, moderately dysregulated, and severely dysregulated profiles in adults with ADHD (Yue et al., 2026). Other person-centered studies have identified profiles differentiated mainly by internalizing psychopathology or neurocognitive indicators, illustrating that findings depend on the measures included (Rodriguez et al., 2026; Cerny et al., 2025). Because these studies differ in age groups, measures, and classification methods, their subgroup proportions are not directly comparable. The present study applies latent class analysis to all 28 clinician-rated WRAADDS items in a large clinical sample of adults with ADHD, without imposing a predefined ED-based grouping of classes.

This study aimed to (I) describe latent WRAADDS symptom profiles in a specialized outpatient cohort of adults with ADHD and report the proportion of participants assigned to profiles with elevated probabilities in emotion-related WRAADDS domains; (II) compare participants assigned to the exploratory ED-related classes with those assigned to the remaining classes on current ADHD symptom scores and retrospectively reported childhood ADHD symptoms from self-report measures, psychological distress, depressive symptoms, functional impairment, alcohol- and drug-related screening scores, and lifetime suicide attempts; and (III) describe documented psychiatric comorbidities and sociodemographic characteristics and examine their associations with the exploratory ED-related grouping.

Sociodemographic variables were selected because sex differences have been reported for ED-related subgroups in adults with ADHD (Hirsch et al., 2019) and because family status, education, and employment reflect psychosocial functioning, which has been associated with deficient emotional self-regulation (Surman et al., 2013); age was included as a basic descriptive characteristic.

## 2. Materials and methods

### 2.1 Participants

Health records from the specialized outpatient clinic for adult ADHD at the Department of Psychiatry and Psychotherapy, University of Leipzig Medical Center (Germany), were reviewed for patients assessed between 2013 and 2023. ADHD diagnoses were established according to ICD-10 criteria within a standardized diagnostic procedure conducted by clinicians experienced in adult ADHD assessment. Diagnostic evaluations were typically performed by two independent clinicians and were based on clinical judgment in combination with information obtained from several validated assessment instruments, including the semi-structured, clinician-rated Wender-Reimherr Adult Attention Deficit Disorder Scale (WRAADDS; Rösler et al., 2008b), the Adult ADHD Self-Report Scale (ASRS-v1.1; Kessler et al., 2005), the Wender Utah Rating Scale–Short Form (WURS-K; Rösler et al., 2008b), and additional self-report measures (Supplementary Table S1). Primary school reports and previous medical or psychological records were routinely requested and considered when available; information from relatives was obtained only in individual cases. The final diagnosis was established by consensus between the two clinicians; in cases of disagreement, the supervising senior physician made the final decision. ADHD diagnoses were thus based on the overall clinical assessment and not on any single instrument or scale score.

Criteria which were required for inclusion were age ≥18 years, a confirmed ADHD diagnosis according to the ICD-10 criteria, and complete WRAADDS data.

Exclusion criteria comprised organic mental disorder; substance intoxication or withdrawal; psychotic disorder, bipolar disorder, severe major depressive episodes; specific personality disorders (e.g., paranoid, schizoid, or borderline personality disorder); persistent personality changes; intellectual disability; and pervasive developmental disorders.

During the study period, a total of 1,204 participants were screened. One participant was excluded due to age below 18 years. Of the remaining participants, 224 did not meet ICD-10 diagnostic criteria for ADHD and were excluded. An additional 102 participants were excluded due to incomplete WRAADDS data. Furthermore, 128 participants were excluded based on predefined psychiatric exclusion criteria. Among participants excluded due to psychiatric comorbidities, the most frequent primary exclusion diagnoses were personality disorders including borderline personality disorder (n=64), severe major depressive episodes (n=41), bipolar disorder (n=12), pervasive developmental disorders (n=7), and psychotic disorders (n=4) (Supplementary Table S2). The final study sample therefore consisted of 749 adults with a confirmed ADHD diagnosis and complete WRAADDS data.

Psychiatric comorbidities analyzed in this study were identified from ICD-10 diagnoses documented in the medical records; no structured diagnostic interview was used.

The study was approved by the Ethics Committee of the Faculty of Medicine of Leipzig University (17523-ek); the requirement for informed consent was waived for this retrospective analysis.

### 2.2 Measures

The assessment battery included ADHD-related and broader clinical measures to characterize complementary aspects of the clinical phenotype. The WRAADDS served as a clinician-rated measure of adult ADHD symptom domains and formed the basis for the latent class analysis. The ASRS-v1.1, ADHD-SR, CAARS, and WURS-K provided complementary self-report perspectives on current symptom severity, ADHD presentation, and retrospective childhood symptoms. Additional instruments assessed functional impairment, psychological distress, depressive symptoms, personality styles, and substance use. Thus, the measures were not intended as redundant diagnostic definitions of ADHD, but to characterize clinical severity, functional impairment, and comorbid symptoms relevant to the study objectives.

Wender-Reimherr Adult Attention Deficit Disorder Scale (WRAADDS; Rösler et al., 2008b): The WRAADDS is a clinician-rated interview comprising 28 items across seven domains: attention difficulties, hyperactivity/restlessness, temper, affective lability, emotional overreactivity, disorganization, and impulsivity. Each item is rated from 0 (absent) to 2 (moderate to severe). According to the scoring guidelines, a domain is considered present when the domain score (the sum of its item ratings) is at least equal to the number of items in the domain and the global rating for that domain (range 0–4) is at least 2 (moderate).

According to the instrument-specific scoring approach, ADHD-related criteria involve the domains of attention difficulties and hyperactivity/restlessness, as well as at least two additional domains. In the present study, however, WRAADDS information was used as part of the standardized clinical assessment and for the latent class analysis, whereas the final ADHD diagnosis was based on the overall clinical evaluation rather than on WRAADDS scores alone.

For the exploratory ED-related grouping, we considered the WRAADDS domains temper, affective lability, and emotional overreactivity. In the original WRAADDS validation, these domains loaded on an emotional factor distinct from an attention/disorganization factor (Marchant et al., 2013). However, a German study grouped emotional overreactivity with inattention and disorganization, whereas temper and affective lability loaded on a separate factor (Rösler et al., 2008a). We therefore treated these domains as an exploratory, construct-related proxy for ED, not as a dedicated ED measure or a validated ED diagnosis.

Conners’ Adult ADHD Rating Scales (CAARS; Christiansen et al., 2014): The CAARS (long version) is a 66-item self-report instrument that assesses ADHD symptoms, emotional instability, and self-concept. It includes four factor-based scales, three symptom scales based on the Diagnostic and Statistical Manual of Mental Disorders, Fourth Edition (DSM-IV), an ADHD index, and an inconsistency index. The items are rated on a 4-point Likert scale (0=not at all to 3=very strongly/often). Raw scores are converted into age- and sex-standardized T-scores; values >65 indicate clinically significant symptoms.

The Adult ADHD Self-Report Scale (ASRS-v1.1; Kessler et al., 2005) is an 18-item self-report measure developed by the WHO. It comprises a six-item screening component (Part A) and 12 additional items (Part B) assessing symptoms of inattention and hyperactivity/impulsivity. A positive screening result is defined as four or more Part A responses meeting the item-specific thresholds. In the present study, total and subscale scores were analyzed as continuous self-report indices of current ADHD symptom burden and were not used for diagnostic classification.

ADHD Self-Report Scale (ADHD-SR; Rösler et al., 2008b): The ADHD-SR is a validated German self-report tool that assesses 18 DSM-based ADHD symptoms in adults. The items are rated on a 4-point scale (0=not at all to 3=very severe), yielding a total score and separate subscale scores for inattention, hyperactivity, and impulsivity.

Wender Utah Rating Scale-Short Form (WURS-K; Rösler et al., 2008b): The WURS-K retrospectively assesses childhood ADHD symptoms using 25 items, of which 21 contribute to the total score; four are control items. A total score ≥30 indicates a possible history of childhood ADHD.

Functional Level Rating Scale (FLRS, Lauth and Minsel, 2014): The FLRS is a component of the standardized Cologne ADHD Test for Adults, which is designed to assess functional impairments commonly associated with ADHD. The FLRS comprises eight two-item subscales assessing everyday functioning in areas such as organization, planning, memory, and social interaction. A separate single item provides a general functional-level rating; higher scores indicate greater impairment.

Symptom Checklist-90-Revised (SCL-90-R; Kliem and Brähler, 2016): The SCL-90-R assesses general psychological distress across 90 items covering nine symptom dimensions. It yields three global indices: the Global Severity Index, the Positive Symptom Distress Index, and the Positive Symptom Total.

Beck Depression Inventory-II (BDI-II; Hautzinger et al., 2006): The BDI-II measures depressive symptom severity across 21 items. Scores of 14–19 indicate mild, 20–28 moderate, and ≥29 severe depressive symptom severity according to the German scoring categories; the BDI-II does not establish a diagnosis.

The Personality Style and Disorder Inventory (PSSI; Kuhl and Kazén, 2009) is a 140-item self-report questionnaire assessing 14 personality styles. Raw scores are transformed into percentiles and T scores; T scores of 60 or higher indicate an above-average expression of a personality style relative to the normative sample and do not constitute a clinical diagnosis. PSSI scores were analyzed exploratorily.

Alcohol Use Disorders Identification Test (AUDIT; Saunders et al., 1993; Dybek et al., 2006): The AUDIT consists of 10 items designed to screen for harmful or hazardous alcohol use. The recommended cutoff scores may vary slightly depending on national drinking norms.

Drug Use Disorders Identification Test (DUDIT; Berman et al., 2003; European Monitoring Centre for Drugs and Drug Addiction, 2020): The DUDIT includes 11 items assessing drug use behavior. A total score ≥2 (women) or ≥6 (men) indicates possible drug-related problems.

### 2.3 Statistical analyses

#### 2.3.1 Latent class analysis

Latent class analyses used R 4.3.3 (R Core Team, 2024) and poLCA 1.6.0.1 (Linzer and Lewis, 2011); all other analyses used R 4.6.1 (R Core Team, 2026). All 28 WRAADDS items from the 749 participants were entered as three-category indicators (1=absent, 2=mild, 3=moderate to severe; i.e., the original 0–2 ratings shifted by one). Models with one to eight classes were estimated using 100 random starts each for one to six classes, 170 starts for seven classes, and 130 starts for eight classes. The selected six-class model was re-estimated with 500 random starts. Model selection considered the Akaike information criterion (AIC), corrected AIC (AICc), Bayesian information criterion (BIC), sample-size-adjusted BIC (SABIC), the bootstrap likelihood ratio test (see below), minimum class size, and the interpretability of the class profiles. Classification quality of the selected model was evaluated separately using relative entropy, average posterior probabilities, and odds of correct classification; these indices were not used to determine the number of classes.

Adjacent models (k−1 vs. k classes) up to six classes were compared using a parametric bootstrap likelihood ratio test with 100 bootstrap samples. To assess whether a higher log-likelihood could be found for the six-class model, we performed a two-stage start-value search: two independent sets of 2,500 random starts were each run for 100 iterations, and the 50 starts with the highest log-likelihood in each set were continued to convergence. Absolute fit of the six-class model was assessed using bivariate Pearson statistics for all 378 item pairs. The pairwise Pearson chi-square contributions were summed to obtain an aggregate statistic, which was compared with a parametric-bootstrap reference distribution based on 100 data sets simulated from the fitted six-class model; the six-class model was refitted to each simulated data set. The pairwise statistics were also summarized separately for item pairs within and between WRAADDS domains.

Class profiles were interpreted using the conditional item response probabilities. The latent class analysis did not estimate an ED grouping, and no validated categorical ED cut-off was available. The grouping used for subsequent comparisons was therefore exploratory and based on post hoc inspection of the class profiles. LC1, LC2, and LC4 each showed mean probabilities of approximately .62 or higher for a moderate-to-severe rating in at least two of the three emotion-related WRAADDS domains (temper, affective lability, and emotional overreactivity). In the remaining classes, the mean probability was below .47 in each of these three domains. On this descriptive basis, LC1, LC2, and LC4 were grouped as ED-related for exploratory comparisons; participants assigned to these classes are hereafter referred to as the ADHD+ED group, and participants assigned to the remaining classes as the ADHD group. These labels denote the exploratory latent class grouping and do not represent a clinical diagnosis or a validated categorical measure of ED. Individuals were assigned to the class with the highest posterior probability (modal assignment). An average posterior probability of .70 or higher is commonly used as a benchmark for adequate class separation, although recommended cutoffs vary (Sorgente et al., 2025). As a sensitivity analysis, the posterior-weighted proportion in the ED-related classes was calculated by summing, for each participant, the posterior probabilities for LC1, LC2, and LC4 and averaging these sums across participants. To assess sensitivity to the grouping decision, this proportion was recalculated under alternative exploratory groupings. In a further sensitivity analysis, the main group comparisons were repeated using the five-class solution, with the ADHD+ED and ADHD groups defined from the class profiles in the three emotion-related WRAADDS domains.

The R scripts used for the latent class analysis and the item-coding information are available at https://doi.org/10.5281/zenodo.23738025.

#### 2.3.2 Analysis of group differences

Group differences between the ADHD+ED and ADHD groups were analyzed in R (version 4.6.1; R Core Team, 2026). Continuous and ordinal variables, including age, number of comorbid diagnoses, the general functional-level rating, and questionnaire scores, were compared using Mann–Whitney U tests with tie-corrected normal approximation. This approach was chosen because several variables were markedly skewed (particularly AUDIT and DUDIT scores) or ordinal and bounded; the same test was used consistently for all continuous and ordinal variables. Effect sizes are reported as *r*. Nominal variables were compared using Fisher’s exact test for 2 × 2 tables or the Fisher–Freeman–Halton test for larger tables, using exact *p*-values where feasible and otherwise Monte Carlo *p*-values based on 100,000 simulations with a fixed seed. Effect sizes are reported as odds ratios or Cramér’s *V*, respectively. WRAADDS domain scores were summarized descriptively because the grouping was derived from these items. Benjamini–Hochberg (BH) correction (Benjamini and Hochberg, 1995) was applied within defined test families (participant characteristics, clinical measures, comorbidity, PSSI, and FLRS); PSSI and FLRS analyses were exploratory. Each comparison used available cases. Documented lifetime suicide attempts were summarized descriptively (Table 5).

Continuous variables are summarized as means ± standard deviations (M ± SD), and categorical variables as counts and percentages, n (%). All tests were two-sided. Statistical significance was set at .05; for analyses with Benjamini–Hochberg correction, significance was determined using the adjusted *p*-values.

#### 2.3.3 Sensitivity and missing-data analyses

To assess whether group differences extended beyond non-emotional ADHD symptom severity, we conducted a sensitivity analysis. The core symptom score was calculated as the mean of the z-standardized WRAADDS domain scores for attention difficulties, hyperactivity/restlessness, disorganization, and impulsivity; the three emotion-related domains were excluded. Outcomes comprised the clinical measures and comorbidity variables included in the group comparisons, including the ADHD-SR and ASRS-v1.1 subscale scores. Exploratory PSSI personality-style scores and FLRS subscale scores were excluded; the general functional-level rating was retained as an outcome. For each continuous or ordinal outcome, including the number of comorbid diagnoses, we estimated unadjusted and core-score-adjusted linear regression models with heteroscedasticity-consistent HC3 standard errors (Long and Ervin, 2000). Estimates are reported in raw units with 95% confidence intervals and as standardized differences in outcome SD units. The presence of any recorded comorbid diagnosis was analyzed using logistic regression, with results reported as odds ratios and 95% confidence intervals. As an additional sensitivity analysis for continuous and ordinal outcomes, we performed a rank-transformed analysis of covariance (ANCOVA; Conover and Iman, 1982) using the ranked outcome and ranked core symptom score, with HC3 standard errors. We also examined partial Spearman correlations between each emotion-related WRAADDS domain and each outcome, controlling for the core symptom score. Benjamini–Hochberg correction was applied within the clinical and comorbidity test families, separately for each analysis; for the dimensional analysis, correction was applied jointly across all outcomes and the three emotion-related domains within each family. Because group assignment, the core symptom score, and the emotion-related domains were all derived from the WRAADDS, these analyses assess associations beyond non-emotional core symptom severity within the same instrument; they do not independently validate ED.

To examine possible bias due to missing self-report data, we compared participants with and without available data for each instrument. Differences in the proportion of missing data between the ADHD+ED and ADHD groups were tested using Fisher’s exact test. Completers and non-completers were compared on variables available for all participants using Mann–Whitney U tests for age, year of assessment, WRAADDS domain scores, and number of comorbid diagnoses, and the Fisher–Freeman– Halton test for gender. Benjamini–Hochberg correction was applied across all missing-data tests and completer–non-completer comparisons.

## 3 Results

### 3.1 Selection of the latent class model

Fit indices for models with one to eight classes are presented in Supplementary Table S3, and class sizes are shown in Supplementary Table S4. The information criteria did not converge on a single solution: BIC and AICc were lowest for four classes, although the BIC differed by less than one point from that for three classes; SABIC was lowest for five classes; and AIC continued to decrease through eight classes. The best log-likelihoods for the seven- and eight-class models were reached in only 2 of 170 and 1 of 130 random starts, respectively, and were therefore not reliably recovered. The bootstrap likelihood ratio test was significant for each comparison from one versus two through five versus six classes (p=.0099 in each case, the smallest attainable value with 100 bootstrap samples) and did not identify an optimal number of classes.

The six-class model showed residual misfit in the aggregate bivariate fit check (1929.1; parametric bootstrap p=.0099). Residual associations were greater within than between WRAADDS domains (mean pairwise statistics 14.7 vs. 3.8), consistent with local dependence among items within domains. In the two-stage start-value search described in Section 2.3.1, the best six-class log-likelihood was reached in 24 of the 100 final runs (7 and 17 in the two sets), and no higher value was found; all 24 reproduced the same person-level class assignments as the reported solution. The other 76 runs were within one log-likelihood unit of the best solution (adjusted Rand indices .85–.97; κ for the ED-related grouping .92–.99).

We retained the six-class solution as the primary solution because, compared with the five-class solution, it provided an interpretable distinction between a profile with globally lower symptom probabilities (PR) and a predominantly inattentive profile (IA), while maintaining adequate classification quality. Average posterior probabilities ranged from .87 to .95, exceeding .70, a commonly used benchmark for adequate class separation, although recommended cutoffs vary (Sorgente et al., 2025). Odds of correct classification ranged from 18.4 to 165.1 (Supplementary Tables S5 and S6). The five-class solution was statistically at least as well supported and was examined in sensitivity analyses. Based on most-likely class assignment, six-class sizes ranged from 9.1% to 27.2% (Table 1); posterior-estimated proportions ranged from 9.3% to 27.2% (Supplementary Table S7).

**Table 1.** Summary of latent class characteristics and rationale for class labels.

| Latent class | n (%) | Descriptive label | Rationale for class label | ED-related pattern | Group for subsequent analyses |
| --- | --- | --- | --- | --- | --- |
| LC1 | 68 (9.1) | HI+ED | Predominant hyperactivity/restlessness and impulsivity with ED-related features | Elevated | ADHD+ED |
| LC2 | 184 (24.6) | IA/HI+ED | Broad elevation across inattention, hyperactivity/restlessness, impulsivity, and ED-related features | Elevated | ADHD+ED |
| LC3 | 79 (10.5) | PR | Lower overall symptom probabilities relative to the other classes, consistent with a partially remitting symptom profile | Not consistently elevated | ADHD |
| LC4 | 204 (27.2) | IA+ED | Predominant inattention with ED-related features | Elevated | ADHD+ED |
| LC5 | 93 (12.4) | IA | Predominant inattention without consistent ED-related features | Not consistently elevated | ADHD |
| LC6 | 121 (16.2) | HI | Predominant hyperactivity/restlessness and impulsivity without consistent ED-related features | Not consistently elevated | ADHD |

**Note.** ADHD, attention-deficit/hyperactivity disorder; ADHD+ED, the exploratory group assigned to classes with elevated probabilities in emotion-related WRAADDS domains; ED, emotional dysregulation; HI, hyperactivity/impulsivity; IA, inattention; LC, latent class; PR, partially remitting symptoms. The ADHD+ED grouping is exploratory and does not represent a clinical diagnosis or validated categorical measure of ED.

Class characteristics and descriptive labels are summarized in Table 1, based on the conditional item response probabilities shown in Supplementary Figure S1; mean probabilities in the emotion-related WRAADDS domains are presented in Supplementary Table S8. Under the primary exploratory grouping, 60.9% of participants were assigned to classes with elevated probabilities in emotion-related domains. Across the alternative groupings, this proportion ranged from 24.6% to 77.0%; the posterior-weighted estimate for the primary grouping was 60.6%, and the five-class grouping yielded 65.2% (Supplementary Table S9).

### 3.2 Demographic and clinical characteristics

The study included 749 adults with ADHD. Demographic and clinical characteristics of the total sample and by group are summarized in Table 2. Age and gender distributions did not differ significantly between the ADHD+ED and ADHD groups. ADHD presentation (ADHD-SR; n=692) differed between groups (Cramér’s *V*=.21, BH-adjusted *p*<.001); the inattentive presentation was less common in the ADHD+ED group.

**Table 2.**
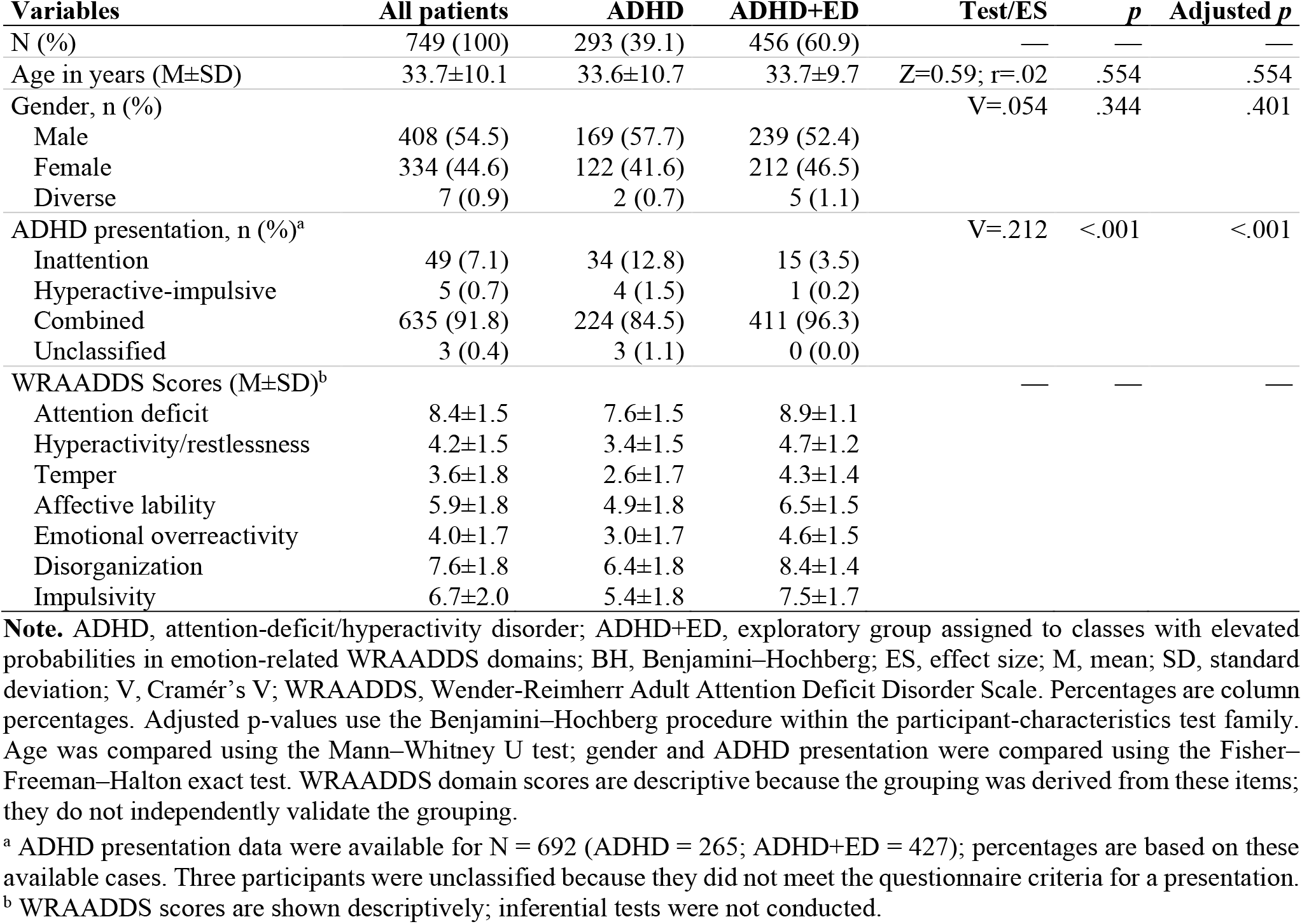
Demographic and clinical characteristics by group.

WRAADDS domain scores were higher in the ADHD+ED group across all seven domains (Table 2). Because the grouping was derived from WRAADDS item-response patterns, these differences are reported descriptively and do not provide independent validation of the grouping.

### 3.3 Sociodemographic characteristics by group

Table 3 summarizes sociodemographic characteristics by group. No significant differences were observed between the ADHD+ED and ADHD groups in family status, highest school qualification, education, or employment status (all BH-adjusted p≥.238). The ADHD+ED group had numerically higher proportions of participants who were single, unemployed, or retired; these descriptive differences were not statistically significant.

**Table 3.** Sociodemographic characteristics by group.

| Variable | All patients | ADHD | ADHD+ED | Test/ES | *p* | Adjusted *p* |
| --- | --- | --- | --- | --- | --- | --- |
| Family status, n (%) |  |  |  | V=.107 | .200 | .350 |
| Single | 237 (31.6) | 83 (28.3) | 154 (33.8) |  |  |  |
| Partnership/married | 374 (49.9) | 146 (49.8) | 228 (50.0) |  |  |  |
| Divorced | 25 (3.3) | 10 (3.4) | 15 (3.3) |  |  |  |
| Widowed | 3 (0.4) | 0 (0.0) | 3 (0.7) |  |  |  |
| Other/unknown | 110 (14.7) | 54 (18.4) | 56 (12.3) |  |  |  |
| Highest school qualification<sup>a</sup>, n (%) |  |  |  | V=.101 | .302 | .401 |
| No formal qualification | 14 (1.9) | 5 (1.7) | 9 (2.0) |  |  |  |
| Secondary school certification | 270 (36.0) | 92 (31.4) | 178 (39.0) |  |  |  |
| University entrance qualification | 421 (56.2) | 180 (61.4) | 241 (52.9) |  |  |  |
| Special needs school certification | 5 (0.7) | 1 (0.3) | 4 (0.9) |  |  |  |
| Other/unknown | 39 (5.2) | 15 (5.1) | 24 (5.3) |  |  |  |
| Education, n (%) |  |  |  | V=.124 | .068 | .238 |
| No education | 83 (11.1) | 28 (9.6) | 55 (12.1) |  |  |  |
| In vocational training | 25 (3.3) | 11 (3.8) | 14 (3.1) |  |  |  |
| In higher education | 125 (16.7) | 55 (18.8) | 70 (15.4) |  |  |  |
| Completed vocational Training | 300 (40.1) | 100 (34.1) | 200 (43.9) |  |  |  |
| Completed university degree | 198 (26.4) | 92 (31.4) | 106 (23.2) |  |  |  |
| Other/unknown | 18 (2.4) | 7 (2.4) | 11 (2.4) |  |  |  |
| Employment status, n (%) |  |  |  | V=.111 | .105 | .244 |
| Unemployed | 126 (16.8) | 43 (14.7) | 83 (18.2) |  |  |  |
| Employed | 365 (48.7) | 148 (50.5) | 217 (47.6) |  |  |  |
| Student or in training | 191 (25.5) | 84 (28.7) | 107 (23.5) |  |  |  |
| Retired | 12 (1.6) | 2 (0.7) | 10 (2.2) |  |  |  |
| Other/unknown | 55 (7.3) | 16 (5.5) | 39 (8.6) |  |  |  |

**Note.** ADHD, attention-deficit/hyperactivity disorder; ADHD+ED, exploratory group assigned to classes with elevated probabilities in emotion-related WRAADDS domains; ES, effect size; V, Cramér's V. Percentages are column percentages. Categories were compared using the Fisher–Freeman–Halton exact test. Adjusted *p*-values use the Benjamini–Hochberg procedure within the participant-characteristics test family. Tests were performed on the original coding categories; some categories are combined for display.
<sup>a</sup> School qualifications follow the German education system. “Secondary school certificate” includes lower and middle school-leaving certificates.

### 3.4 Comorbid disorders by group

Table 4 summarizes documented comorbid diagnoses by group. The proportion of participants with any recorded comorbid diagnosis did not differ between groups, whereas the ADHD+ED group had a higher number of comorbid diagnoses and a different distribution across comorbidity categories. Descriptively, the multiple-disorders and substance-use-disorder categories were more frequent in the ADHD+ED group, whereas depressive and adjustment disorder categories were more frequent in the ADHD group. Participants with more than one comorbid diagnosis were assigned to the multiple-disorders category; frequencies in the individual disorder categories should therefore be interpreted in light of this classification. On self-report measures, the ADHD+ED group had higher depressive symptom scores on the BDI-II and higher drug-use screening scores on the DUDIT, whereas alcohol-use screening scores on the AUDIT did not differ.

**Table 4.**
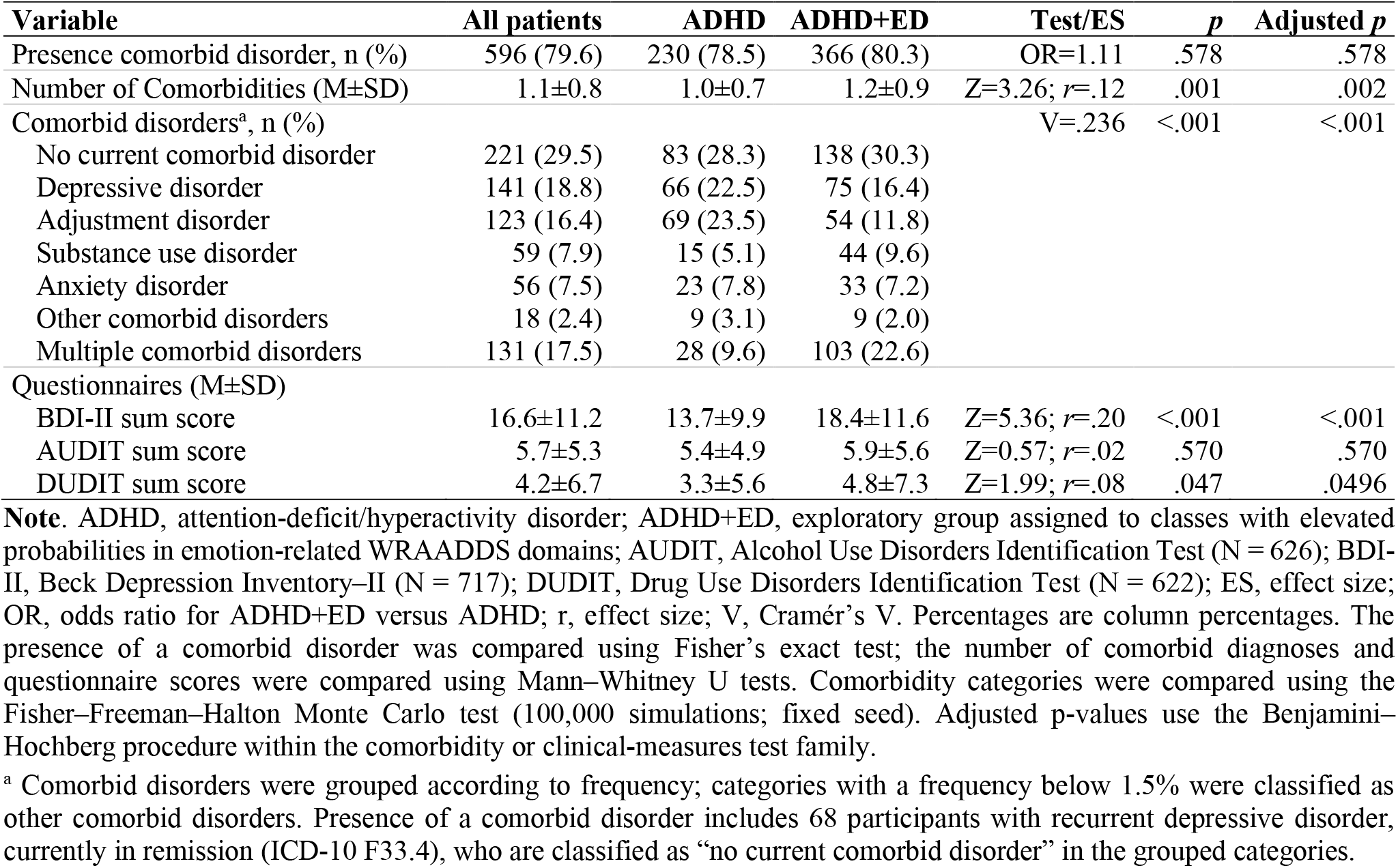
Psychiatric comorbidities and related clinical measures by group.

In exploratory PSSI analyses, BH-adjusted group differences were observed for 12 of the 14 personality-style scales: scores were higher in the ADHD+ED group for 11 scales and lower for the obsessive-compulsive style, whereas schizoid and rhapsodic scores did not differ (Supplementary Table S10). No group mean reached a T-score of 60. These exploratory findings do not establish distinct personality profiles.

### 3.5 Symptom severity and functional impairment by group

Table 5 summarizes group comparisons in ADHD symptom measures, psychological distress, and functional impairment. The ADHD+ED group had higher scores on all CAARS scales, including more problems with self-concept, and on the ADHD-SR, ASRS-v1.1, and WURS-K (all BH-adjusted *p*<.001), with small-to-moderate effect sizes. Because the latent classes were derived from WRAADDS profiles that include core ADHD symptom domains as well as emotion-related domains, these differences were partly expected; their robustness after adjustment for core symptom severity is reported in Section 3.6.

**Table 5.**
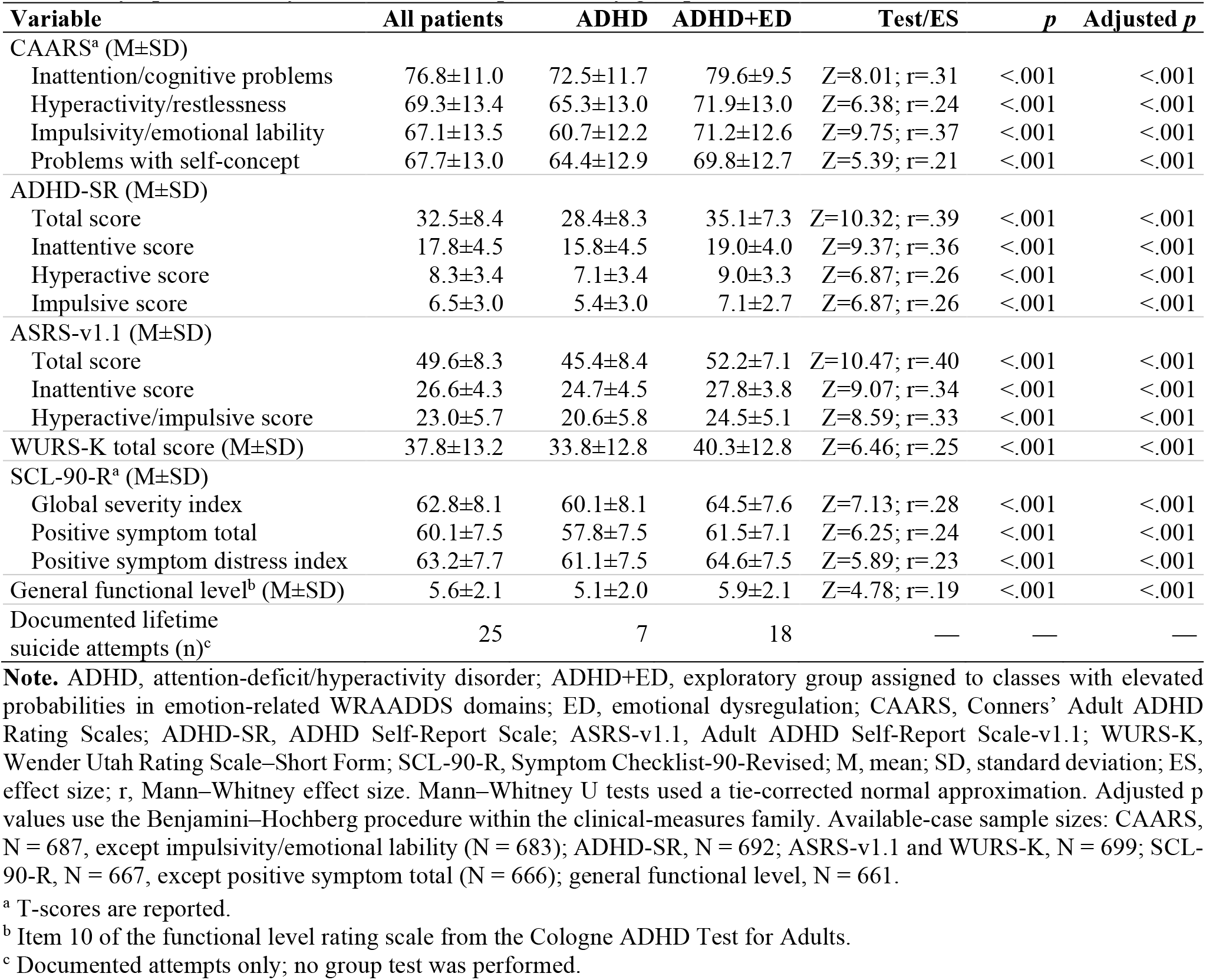
Symptom severity and functional impairment by group.

| Variable | All patients | ADHD | ADHD+ED | Test/ES | *p* | Adjusted *p* |
| --- | --- | --- | --- | --- | --- | --- |
| CAARS<sup>a</sup> (M±SD) |  |  |  |  |  |  |
| Inattention/cognitive problems | 76.8±11.0 | 72.5±11.7 | 79.6±9.5 | Z=8.01; *r*=.31 | <.001 | <.001 |
| Hyperactivity/restlessness | 69.3±13.4 | 65.3±13.0 | 71.9±13.0 | Z=6.38; *r*=.24 | <.001 | <.001 |
| Impulsivity/emotional lability | 67.1±13.5 | 60.7±12.2 | 71.2±12.6 | Z=9.75; *r*=.37 | <.001 | <.001 |
| Problems with self-concept | 67.7±13.0 | 64.4±12.9 | 69.8±12.7 | Z=5.39; *r*=.21 | <.001 | <.001 |
| ADHD-SR (M±SD) |  |  |  |  |  |  |
| Total score | 32.5±8.4 | 28.4±8.3 | 35.1±7.3 | Z=10.32; *r*=.39 | <.001 | <.001 |
| Inattentive score | 17.8±4.5 | 15.8±4.5 | 19.0±4.0 | Z=9.37; *r*=.36 | <.001 | <.001 |
| Hyperactive score | 8.3±3.4 | 7.1±3.4 | 9.0±3.3 | Z=6.87; *r*=.26 | <.001 | <.001 |
| Impulsive score | 6.5±3.0 | 5.4±3.0 | 7.1±2.7 | Z=6.87; *r*=.26 | <.001 | <.001 |
| ASRS-v1.1 (M±SD) |  |  |  |  |  |  |
| Total score | 49.6±8.3 | 45.4±8.4 | 52.2±7.1 | Z=10.47; *r*=.40 | <.001 | <.001 |
| Inattentive score | 26.6±4.3 | 24.7±4.5 | 27.8±3.8 | Z=9.07; *r*=.34 | <.001 | <.001 |
| Hyperactive/impulsive score | 23.0±5.7 | 20.6±5.8 | 24.5±5.1 | Z=8.59; *r*=.33 | <.001 | <.001 |
| WURS-K total score (M±SD) | 37.8±13.2 | 33.8±12.8 | 40.3±12.8 | Z=6.46; *r*=.25 | <.001 | <.001 |
| SCL-90-R<sup>a</sup> (M±SD) |  |  |  |  |  |  |
| Global severity index | 62.8±8.1 | 60.1±8.1 | 64.5±7.6 | Z=7.13; *r*=.28 | <.001 | <.001 |
| Positive symptom total | 60.1±7.5 | 57.8±7.5 | 61.5±7.1 | Z=6.25; *r*=.24 | <.001 | <.001 |
| Positive symptom distress index | 63.2±7.7 | 61.1±7.5 | 64.6±7.5 | Z=5.89; *r*=.23 | <.001 | <.001 |
| General functional level<sup>b</sup> (M±SD) | 5.6±2.1 | 5.1±2.0 | 5.9±2.1 | Z=4.78; *r*=.19 | <.001 | <.001 |
| Documented lifetime suicide attempts (n)<sup>c</sup> | 25 | 7 | 18 | — | — | — |

**Note.** ADHD, attention-deficit/hyperactivity disorder; ADHD+ED, exploratory group assigned to classes with elevated probabilities in emotion-related WRAADDs domains; ED, emotional dysregulation; CAARS, Conners' Adult ADHD Rating Scales; ADHD-SR, ADHD Self-Report Scale; ASRS-v1.1, Adult ADHD Self-Report Scale-v1.1; WURS-K, Wender Utah Rating Scale–Short Form; SCL-90-R, Symptom Checklist-90-Revised; M, mean; SD, standard deviation; ES, effect size; *r*, Mann–Whitney effect size. Mann–Whitney U tests used a tie-corrected normal approximation. Adjusted *p* values use the Benjamini–Hochberg procedure within the clinical-measures family. Available-case sample sizes: CAARS, *N* = 687, except impulsivity/emotional lability (*N* = 683); ADHD-SR, *N* = 692; ASRS-v1.1 and WURS-K, *N* = 699; SCL-90-R, *N* = 667, except positive symptom total (*N* = 666); general functional level, *N* = 661.
<sup>a</sup> T-scores are reported.
<sup>b</sup> Item 10 of the functional level rating scale from the Cologne ADHD Test for Adults.
<sup>c</sup> Documented attempts only; no group test was performed.

The ADHD+ED group also had higher scores across the SCL-90-R global indices, indicating greater psychological distress, and a higher general functional-level rating, indicating greater impairment. Exploratory analyses showed higher impairment scores across all eight FLRS subscales (Supplementary Table S11). Documented lifetime suicide attempts are shown descriptively in Table 5.

### 3.6 Sensitivity and missing-data analyses

In the five-class sensitivity analysis, continuous group differences were unchanged in direction, and categorical distributions were similar to those in the six-class analysis. After BH correction, only the DUDIT total score was no longer statistically significant (Supplementary Table S12).

Self-report data were missing for 4.3% (BDI-II) to 17.0% (DUDIT) of participants; 481 participants (64.2%) had complete data across all self-report instruments (Supplementary Table S13). After BH correction, missingness did not differ between the ADHD+ED and ADHD groups for any instrument. Completer–non-completer comparisons are summarized in the note to Supplementary Table S13.

The core symptom score was associated with group membership (point-biserial *r*=.70). After adjustment for the core symptom score, group differences remained significant for psychological distress, depressive symptoms, functional level, and the CAARS scales for inattention/cognitive problems, problems with self-concept, and impulsivity/emotional lability, although some effects were attenuated. Differences in ADHD-SR and ASRS-v1.1 total scores and subscales, WURS-K scores, DUDIT scores, and the number of comorbid diagnoses were no longer significant in the core-adjusted linear models. The rank-transformed ANCOVA yielded a borderline BH-adjusted result for ADHD-SR inattention (adjusted *p*=.049), which was not significant in the core-adjusted linear model. AUDIT scores and the presence of a comorbid disorder did not differ in either model. For CAARS hyperactivity/restlessness, the adjusted estimate changed direction, but neither the core-adjusted linear model nor the rank-transformed ANCOVA showed a significant difference after BH correction; we therefore do not interpret this as evidence of lower hyperactivity/restlessness in the ADHD+ED group.

In the dimensional analysis, partial Spearman correlations controlling for the core symptom score showed associations of all three emotion-related domains with SCL-90-R global indices and BDI-II scores. Affective lability and emotional overreactivity were also associated with the general functional-level rating and CAARS problems with self-concept. Other domain-specific associations are reported in Supplementary Table S14. Associations with comorbidity measures were not significant; associations with AUDIT and DUDIT scores were small and inconsistent in direction.

### 3.7 Internal consistency

Internal consistency was acceptable to excellent for the CAARS factor scales, ADHD-SR, ASRS-v1.1, WURS-K, BDI-II, AUDIT, DUDIT, and SCL-90-R (Cronbach’s α=.70–.97). It was lower for the exploratory two-item FLRS subscales (α=.48–.78) and low to moderate for the WRAADDS domains (α=.44–.75; Supplementary Table S15).

## 4 Discussion

This retrospective study examined latent symptom profiles in a specialized outpatient sample of adults with ADHD and explored their associations with clinical measures, documented psychiatric comorbidities, and sociodemographic characteristics. For the group comparisons, participants assigned to classes with elevated probabilities in the emotion-related WRAADDS domains were referred to as the ADHD+ED group; those assigned to the remaining classes were referred to as the ADHD group. These labels refer to an exploratory WRAADDS-based grouping, not to a clinical diagnosis or validated categorical measure; accordingly, the findings concern this grouping rather than ED as a broader, dimensional construct.

Latent class analysis of all 28 WRAADDS items yielded several plausible solutions. We retained the six-class solution because it distinguished a partially remitting from a predominantly inattentive profile. In the five-class sensitivity analysis, the direction of continuous group differences was unchanged and categorical distributions were similar. The resulting profiles varied in overall symptom level and in the emotion-related domains: elevated probabilities in these domains occurred alongside hyperactive/impulsive, combined, or inattentive symptom patterns (LC1, LC2, and LC4), but not in the remaining classes. This pattern is broadly consistent with WRAADDS-based studies in adults that distinguished inattentive from emotional dysregulation presentations (Reimherr et al., 2015, 2020) and with a latent class analysis in children in which ED co-occurred with different core-symptom profiles (Yue et al., 2022). The co-occurrence of emotion-related elevations and differences in non-emotional domains underscores the potential overlap between emotional features and overall ADHD symptom severity in these class profiles.

Under the primary grouping, 60.9% of participants were assigned to the ED-related classes. This proportion is not an estimate of ED prevalence: it depended on the exploratory grouping decision, ranged from 24.6% to 77.0% across alternative groupings (Supplementary Table S9), and was based on a WRAADDS-derived proxy rather than a dedicated ED measure. It falls numerically within the 34%–70% range reported for impairing ED in clinic-based samples of adults with ADHD, although the estimates are not directly comparable because the definitions and measures differed (Shaw et al., 2014). The specialized, treatment-seeking setting may have enriched the sample for clinically complex presentations, whereas excluding patients with borderline personality disorder, bipolar disorder, or severe depressive episodes may have lowered the observed proportion. The net effect of these sample-selection factors cannot be determined.

The ADHD+ED and ADHD groups did not differ significantly in age, gender distribution, family status, school qualification, education, or employment status. Although we found no gender difference, Hirsch et al. (2019) reported that women were overrepresented in one ED-related cluster, and a scoping review identified biological sex as a factor associated with ED (Bodalski et al., 2023). Differences in the WRAADDS-based operationalization of ED and in sample composition may contribute to these differing findings. The absence of significant differences in education or employment does not rule out previously reported associations between emotional impulsiveness or deficient emotional self-regulation and impairment in major life activities and psychosocial functioning (Barkley and Fischer, 2010; Surman et al., 2013); our data neither establish nor exclude such associations.

The ADHD+ED group had a higher unadjusted number of documented comorbid diagnoses and was more often assigned to the category of multiple comorbid disorders, consistent with previous adult ADHD studies reporting greater comorbidity in ED-related groups (Hirsch et al., 2019; Brancati et al., 2026). However, the difference in the number of diagnoses was no longer significant after adjustment for core symptom severity. Because participants with more than one diagnosis were assigned to the multiple-disorders category, the higher frequency of single depressive or adjustment disorders in the ADHD group partly reflects this classification and does not indicate a lower burden of these disorders in the ADHD+ED group. On the self-report screening measures, higher BDI-II scores in the ADHD+ED group remained significant after adjustment. A multicenter study using elevated CAARS emotional-lability scores to define ED also reported higher BDI-II scores among adults with ADHD and ED (Huang et al., 2024). The small DUDIT difference was not robust in the five-class or core-adjusted analyses, and AUDIT scores did not differ. These screening scores reflect symptom levels rather than diagnoses; therefore, the findings do not support firm conclusions about substance use or the number of comorbid disorders.

In adults with ADHD, emotional dysregulation was associated with suicidal ideation in a case-control study (Ekici et al., 2026). A clinical study found no significant associations between lifetime suicide attempts and ADHD-related features such as mood swings or anger outbursts (Perotti et al., 2025). In the present study, lifetime suicide attempts were documented in 25 participants and summarized descriptively; no conclusions about group differences can therefore be drawn. Prospective studies with systematic assessment of suicidal ideation and behavior are needed to examine whether the exploratory ADHD+ED grouping is associated with suicide risk.

Treatment effects were not examined in the present study. Reimherr et al. (2015) reported similar improvement during methylphenidate treatment in adults classified as having an ADHD emotional dysregulation presentation and those with an ADHD inattentive presentation. Meta-analyses suggest that nonpharmacological interventions may improve emotional symptoms in children and adults with ADHD (Guo et al., 2022), while psychotherapies may reduce comorbid internalizing symptoms in adults with ADHD (López-Pinar et al., 2020).

Overall, participants assigned to the ADHD+ED group had higher scores on ADHD symptom measures and greater psychological distress, depressive symptoms, and functional impairment. Because group assignment and the core symptom score were both derived from the WRAADDS, comparisons involving WRAADDS domains are partly circular. After adjustment for the core symptom score, differences in the broader self-report ADHD measures (ADHD-SR, ASRS-v1.1, and WURS-K) were no longer significant, whereas differences in psychological distress, depressive symptoms, general functional level, and several CAARS scales remained. The CAARS impulsivity/emotional lability scale overlaps in content with the emotion-related WRAADDS domains and therefore does not provide independent confirmation. In the dimensional analyses, the emotion-related WRAADDS domains remained associated with psychological distress and depressive symptoms after controlling for core symptom severity. The association between emotional impulsiveness and impairment beyond ADHD symptom severity reported by Barkley and Fischer (2010) provides a relevant comparison. However, the cross-sectional design and shared WRAADDS basis do not permit conclusions about direction or an independent effect of ED. The findings suggest that emotion-related features may help characterize clinical heterogeneity among adults with ADHD, but do not establish a distinct clinical category.

### 4.1 Limitations

Despite the strengths of this study, several limitations should be considered. First, the retrospective cross-sectional design does not permit causal inferences regarding the associations between the ED-related grouping, ADHD symptom measures, and psychiatric comorbidities.

Second, the sample was drawn from a single specialized outpatient clinic, and the treatment-seeking population may have included more severe or complex clinical presentations than adults with ADHD in the general population. Patients with specific personality disorders (including borderline personality disorder), bipolar disorder, or current severe depressive episodes were excluded, which limits the generalizability of the findings to adults with ADHD and these comorbidities. Harmful substance use and substance dependence were not exclusion criteria, whereas acute intoxication and withdrawal were. The proportion assigned to the ED-related classes and the group differences should therefore be interpreted as specific to this selected outpatient sample.

Third, comorbid diagnoses were derived from clinical records rather than structured diagnostic interviews, and information from relatives was obtained only in individual cases; the completeness and reliability of documented comorbidity may therefore have varied.

Fourth, ED was operationalized through an exploratory, WRAADDS-based approach rather than a dedicated ED instrument. The grouping therefore relied on the same assessment used to characterize ADHD symptoms, which limits the independence of the ED-related grouping from ADHD symptom severity. Differences in WRAADDS domain scores between the ADHD+ED and ADHD groups are expected consequences of the grouping rather than evidence of its validity. ED is increasingly conceptualized as a dimensional and transdiagnostic construct; estimates may differ when independent ED measures or predefined operational definitions are used.

Fifth, several analytic decisions limit the robustness of the latent class results. The information criteria did not converge on a single solution, and the six-class solution was selected partly on interpretability. Grouping three classes as ED-related was a post hoc decision based on class profiles, and the proportion assigned to these classes varied considerably across alternative groupings. The six-class model showed residual local dependence among items within WRAADDS domains. The ED-related classes also showed higher probabilities in non-emotional domains; adjustment for a core symptom score addressed this only partly, because the score was derived from the same instrument. Group comparisons were based on modal class assignment and therefore did not account for classification uncertainty. The posterior-weighted proportion assigned to the ED-related classes (60.6%) was similar to the proportion based on modal assignment (60.9%); however, the impact of classification uncertainty on the group comparisons was not formally examined.

Sixth, the internal consistency of the WRAADDS domain scores was low to moderate (α = .44–.75), including the emotion-related domains, and interrater reliability of the WRAADDS could not be assessed in this routine clinical sample. This may have reduced the precision of domain-based summaries and of the core symptom score, although the latent class analysis was based on item-level responses.

Seventh, analyses of self-report measures used instrument-specific available-case samples. Missingness was related primarily to the year of assessment, for reasons that could not be determined retrospectively. After correction for multiple testing, missingness did not differ significantly between the ADHD+ED and ADHD groups, although it was numerically more frequent in the ADHD+ED group for the AUDIT and DUDIT. Completers and non-completers also differed in year of assessment and, for some instruments, in WRAADDS domain scores or gender. Bias due to missing data therefore cannot be ruled out, particularly for the AUDIT and DUDIT.

Finally, suicidality was represented only by documented lifetime suicide attempts, which were summarized descriptively. Future research should use prospective, multicenter designs in more diverse clinical and community samples, with independent ED measures and systematic assessment of suicidal ideation and behavior.

### 5 Conclusion

In this specialized outpatient sample of adults with ADHD, 60.9% of participants were assigned to latent classes with elevated probabilities in emotion-related WRAADDS domains; this proportion depended on the exploratory grouping and should not be interpreted as a prevalence estimate of ED. Participants assigned to these classes showed greater psychological distress, depressive symptoms, and functional impairment, and these differences persisted after adjustment for core ADHD symptom severity. Differences in broader self-report ADHD symptom measures and in the number of comorbid diagnoses were not robust to this adjustment.

These findings suggest that emotion-related features are associated with clinical heterogeneity among adults with ADHD in specialized care, but they do not establish a distinct clinical category or causal relationships. Future research should use independent, dimensional measures of ED, longitudinal designs, and studies of interventions targeting emotion-regulation difficulties in adults with ADHD.

## Abbreviations

ADHD: Attention-deficit/hyperactivity disorder
ADHD+ED: Exploratory group assigned to latent classes with elevated probabilities in emotion-related WRAADDS domains
ADHD-SR: ADHD Self-Report Scale
AIC: Akaike information criterion
AICc: Corrected Akaike information criterion
ASRS-v1.1: Adult ADHD Self-Report Scale, version 1.1
AUDIT: Alcohol Use Disorders Identification Test
BDI-II: Beck Depression Inventory–II
BH: Benjamini–Hochberg
BIC: Bayesian information criterion
CAARS: Conners’ Adult ADHD Rating Scales
DSM-IV: Diagnostic and Statistical Manual of Mental Disorders, Fourth Edition
DSM-5-TR: Diagnostic and Statistical Manual of Mental Disorders, Fifth Edition, Text Revision
DUDIT: Drug Use Disorders Identification Test
ED: Emotional dysregulation
FLRS: Functional Level Rating Scale
ICD-10: International Statistical Classification of Diseases and Related Health Problems, 10th Revision
PSSI: Personality Style and Disorder Inventory
SABIC: Sample-size-adjusted Bayesian information criterion
SCL-90-R: Symptom Checklist-90-Revised
WRAADDS: Wender-Reimherr Adult Attention Deficit Disorder Scale
WURS-K: Wender Utah Rating Scale–Short Form

## Author Contributions

MS and NM contributed to the conceptualization, study design, and project administration. TT and CM were responsible for data collection and curation. NM and JH conducted the data analysis and contributed to the interpretation of the results, including the preparation of figures and tables. TT and NM drafted the original manuscript and approved the final version. All the authors read and approved the final manuscript.

## Acknowledgments

R code was reviewed and refined using a local LLM (vllm-llama-4-Scout-17b-16e-instruct) via BIA-Bob (version 0.36.4) and the beta AI assistant in RStudio (Posit). Claude Opus 5.5 (Anthropic; claude.ai, October 2026) assisted with language editing of the manuscript and reviewer responses.

The authors reviewed all AI-assisted outputs and take responsibility for the analyses and final manuscript.

## Data Availability Statement

The datasets presented in this article are not publicly available because they contain sensitive clinical information collected in routine care. Requests to access the datasets may be directed to the corresponding author and will be considered in accordance with applicable data protection regulations. The R scripts used for the latent class analysis and the item-coding information are available in the GitHub repository and archived on Zenodo as a citable, version-specific record (DOI: https://doi.org/10.5281/zenodo.23738025).

## Ethics statement

The study was conducted in accordance with the latest version of the Declaration of Helsinki and was reviewed by the Ethics Committee of the Faculty of Medicine of Leipzig University (Ethics Audit Number: 17523-ek). In accordance with applicable laws, we have taken the necessary steps to ensure that the rights of the participants are protected. In accordance with the provisions of Articles 89 and 27 of the General Data Protection Regulation and Section 27 of the German Data Protection Act, we have waived the requirement for retrospective informed consent.

## Supplementary material

Supplementary Tables S1–S15 and Figure S1 are provided in the Supplementary Material file.

