## Supplementary Material (Tables S1 to S15, Figure S1) for "Exploratory Symptom Profiles and Clinical Correlates in Adults with Attention-Deficit/Hyperactivity Disorder: A Retrospective Study Using the Wender-Reimherr Adult Attention Deficit Disorder Scale"

### *Participants*

**Table S1**. Self-report instruments used in the study.

| Instrument | Construct assessed | Brief description | Reference |
|---|---|---|---|
| CAARS | ADHD symptom severity | Self-report assessment of adult ADHD symptoms, including inattention, hyperactivity/restlessness, impulsivity/emotional lability, and self-concept | (Christiansen et al. 2014) |
| ASRS-v1.1 | ADHD screening and symptom severity | WHO-developed self-report screening tool for adult ADHD symptoms | (Kessler et al. 2005) |
| ADHD-SR | ADHD symptom severity | German self-report scale assessing DSM-based ADHD symptoms in adults | (Rösler et al. 2008) |
| WURS-K | Childhood ADHD symptoms | Retrospective self-report assessment of childhood ADHD symptoms | (Rösler et al. 2008) |
| FLRS | Functional impairment | Self-report measure of functional impairments associated with adult ADHD | (Lauth and Minsel 2014) |
| SCL-90-R | General psychological distress | Self-report assessment of global psychopathological distress and symptom dimensions | (Kliem and Brähler 2016) |
| BDI-II | Depressive symptoms | Self-report measure of depressive symptom severity | (Hautzinger et al. 2006) |
| PSSI | Personality styles | Self-report assessment of personality styles and traits | (Kuhl and Kazén 2009) |
| AUDIT | Alcohol use | Screening questionnaire for harmful or hazardous alcohol consumption | (Saunders et al. 1993; Dybek et al. 2006) |
| DUDIT | Drug use | Screening questionnaire for drug-related problems | (Berman et al. 2003; European Monitoring Centre for Drugs and Drug Addiction (EMCDDA) 2020) |

**Note.** ADHD, attention-deficit/hyperactivity disorder; ADHD-SR, ADHD Self-Report Scale; ASRS-v1.1, Adult ADHD Self-Report Scale version 1.1; AUDIT, Alcohol Use Disorders Identification Test; BDI-II, Beck Depression Inventory-II; CAARS, Conners' Adult ADHD Rating Scales; DSM, Diagnostic and Statistical Manual of Mental Disorders; DUDIT, Drug Use Disorders Identification Test; FLRS, Functional Level Rating Scale; PSSI, Personality Style and Disorder Inventory; SCL-90-R, Symptom Checklist-90-Revised; WHO, World Health Organization; WURS-K, Wender Utah Rating Scale–Short Form.

**Table S2**. Psychiatric comorbidities leading to exclusion.

| Primary exclusion diagnosis | n |
|---|---|
| Specific personality disorders (e.g., paranoid, schizoid, or borderline) | 64 |
| Severe major depressive episode | 41 |
| Bipolar disorder | 12 |
| Pervasive developmental disorder | 7 |
| Psychotic disorder | 4 |
| **Total** | **128** |

**Note.** Patients were assigned to one primary exclusion category based on the main clinical diagnosis leading to exclusion.

### *Selection of the latent class model*

**Table S3**. Fit indices and classification diagnostics for one- to eight-class latent class models.

| Classes | LL | Free parameters | AIC | AICc | BIC | SABIC | Relative entropy | Smallest class, n (%) | BLRT LR (p) |
|---|---|---|---|---|---|---|---|---|---|
| 1 | −18481.4 | 56 | 37074.8 | 37084.0 | 37333.4 | 37155.6 | — | 749 (100.0) | — |
| 2 | −17635.4 | 113 | 35496.7 | 35537.3 | 36018.6 | 35659.8 | .815 | 319 (42.6) | 1692.1 (.0099) |
| 3 | −17431.6 | 170 | 35203.3 | 35303.9 | 35988.5 | 35448.6 | .807 | 123 (16.4) | 407.4 (.0099) |
| 4 | −17242.6 | 227 | 34939.1 | **35137.8** | **35987.6** | 35266.8 | .800 | 155 (20.7) | 378.1 (.0099) |
| 5 | −17119.0 | 284 | 34806.1 | 35155.0 | 36117.8 | **35216.0** | .826 | 71 (9.5) | 247.0 (.0099) |
| 6 | −17035.4 | 341 | 34752.9 | 35325.9 | 36327.8 | 35245.0 | .836 | 68 (9.1) | 167.2 (.0099) |
| 7[a] | −16952.8 | 398 | 34701.7 | 35609.1 | 36539.9 | 35276.1 | .852 | 71 (9.5) | — |
| 8[a] | −16873.0 | 455 | **34656.0** | 36072.2 | 36757.5 | 35312.7 | .868 | 50 (6.7) | — |

**Note.** LL, log-likelihood; AIC, Akaike information criterion; AICc, corrected AIC; BIC, Bayesian information criterion; SABIC, sample-size-adjusted BIC; BLRT, bootstrap likelihood ratio test. Minimum values are shown in bold; the BIC for four classes (35987.6) was less than one point lower than that for three classes (35988.5). BLRTs were conducted for comparisons from one versus two through five versus six classes, with 100 bootstrap samples; .0099 is the smallest attainable p-value. For the six-class model, the separate 500-start run reached the best log-likelihood once; in the additional two-stage search, the best log-likelihood was reached in 24 of 100 continued runs, and no higher value was found. Aggregate bivariate Pearson statistic for the six-class model: 1929.1 (37 of 378 item pairs exceeded 9.49, the .05 critical value of $\chi^2$ with 4 df); parametric bootstrap (100 simulated data sets): mean 1161.4 (SD 40.0), 95th percentile 1223.7, maximum 1254.9; p=.0099.

[a] The best log-likelihood was reached in only 2 of 170 starts for the seven-class model and 1 of 130 starts for the eight-class model; these solutions were not reliably recovered.

**Table S4**. Class sizes and estimated proportions by model.

**A.** Observed class counts based on most-likely class assignment.

| Model | Counts by class |
|---|---|
| 2-class | LC1: 430; LC2: 319 |
| 3-class | LC1: 123; LC2: 281; LC3: 345 |
| 4-class | LC1: 190; LC2: 155; LC3: 161; LC4: 243 |
| 5-class | LC1: 123; LC2: 138; LC3: 219; LC4: 71; LC5: 198 |
| 6-class | LC1: 68; LC2: 184; LC3: 79; LC4: 204; LC5: 93; LC6: 121 |
| 7-class | LC1: 143; LC2: 175; LC3: 74; LC4: 71; LC5: 97; LC6: 111; LC7: 78 |
| 8-class | LC1: 68; LC2: 71; LC3: 165; LC4: 84; LC5: 76; LC6: 168; LC7: 50; LC8: 67 |

**B**. Model-estimated class proportions.

| Model | Estimated proportions by class |
|---|---|
| 2-class | LC1: .57075; LC2: .42925 |
| 3-class | LC1: .17011; LC2: .37118; LC3: .45871 |
| 4-class | LC1: .25112; LC2: .21278; LC3: .21686; LC4: .31924 |
| 5-class | LC1: .16923; LC2: .18633; LC3: .28588; LC4: .09424; LC5: .26432 |
| 6-class | LC1: .09304; LC2: .24066; LC3: .10609; LC4: .27224; LC5: .12278; LC6: .16518 |
| 7-class | LC1: .19452; LC2: .23358; LC3: .10070; LC4: .09537; LC5: .12428; LC6: .14763; LC7: .10392 |
| 8-class | LC1: .08805; LC2: .09330; LC3: .22082; LC4: .11207; LC5: .10257; LC6: .22292; LC7: .06814; LC8: .09214 |

**Note.** Counts are based on assignment to the class with the highest posterior probability. Estimated class proportions are model-estimated class-membership proportions and may therefore differ from observed counts divided by the sample size (N=749). LC, latent class.

**Table S5**. Average posterior probabilities by assigned class in the six-class solution.

| | LC1 | LC2 | LC3 | LC4 | LC5 | LC6 |
|---|---|---|---|---|---|---|
| LC1 | **.895** | .035 | .003 | .024 | .000 | .043 |
| LC2 | .027 | **.903** | .000 | .062 | .000 | .009 |
| LC3 | .002 | .000 | **.951** | .013 | .012 | .021 |
| LC4 | .009 | .049 | .002 | **.873** | .021 | .045 |
| LC5 | .001 | .000 | .023 | .057 | **.883** | .036 |
| LC6 | .015 | .014 | .013 | .053 | .038 | **.867** |

**Note.** Rows indicate assignment to the most likely class; columns indicate the average posterior probability of membership in each latent class. Diagonal values are the average posterior probabilities for the assigned classes (AvePP). LC, latent class.

**Table S6**. Odds of correct classification for the six-class solution.

| Latent class | Descriptive label | OCC |
|---|---|---|
| LC1 | HI+ED | 83.2 |
| LC2 | IA/HI+ED | 29.2 |
| LC3 | PR | 165.1 |
| LC4 | IA+ED | 18.4 |
| LC5 | IA | 53.9 |
| LC6 | HI | 33.0 |

**Note.** OCC, odds of correct classification; LC, latent class; HI, hyperactivity/impulsivity; IA, inattention; ED, emotional dysregulation; PR, partially remitting symptoms.

**Table S7**. Observed and posterior-estimated class sizes for the six-class solution.

| | Observed n (most likely class) | Expected n (sum of posterior probabilities) | Estimated proportion |
|---|---|---|---|
| LC1 | 68 | 69.7 | .093 |
| LC2 | 184 | 180.3 | .241 |
| LC3 | 79 | 79.5 | .106 |
| LC4 | 204 | 203.9 | .272 |
| LC5 | 93 | 92.0 | .123 |
| LC6 | 121 | 123.7 | .165 |

**Note.** Observed counts are based on assignment to the class with the highest posterior probability. Expected counts are sums of individual posterior probabilities for each class. Estimated proportions are model-based class-membership proportions. LC, latent class.

**Table S8**. Mean conditional probabilities of moderate-to-severe ratings in the emotion-related WRAADDS domains by latent class.

| Latent class | n (%) | Descriptive label | Temper (3 items) | Affective lability (4 items) | Emotional overreactivity (3 items) | Grouped as ED-related for exploratory comparisons |
|---|---|---|---|---|---|---|
| LC1 | 68 (9.1) | HI+ED | .72 | .63 | .18 | Yes |
| LC2 | 184 (24.6) | IA/HI+ED | .76 | .80 | .75 | Yes |
| LC3 | 79 (10.5) | PR | .13 | .34 | .05 | No |
| LC4 | 204 (27.2) | IA+ED | .25 | .62 | .65 | Yes |
| LC5 | 93 (12.4) | IA | .08 | .40 | .39 | No |
| LC6 | 121 (16.2) | HI | .39 | .46 | .36 | No |

**Note.** Values are mean conditional probabilities of a category 3 rating (moderate to severe), averaged across items within each domain; values are rounded to two decimal places. Items were coded 1=absent, 2=mild, and 3=moderate to severe, corresponding to the original 0–2 ratings shifted by one. Class sizes are based on assignment to the most likely class (N=749). The ED-related grouping is an exploratory, profile-based classification for subsequent comparisons; it is not a clinical diagnosis or a validated categorical measure of emotional dysregulation. ED, emotional dysregulation; HI, hyperactivity/impulsivity; IA, inattention; LC, latent class; PR, partially remitting symptoms; WRAADDS, Wender-Reimherr Adult Attention Deficit Disorder Scale.

**Table S9.** Sensitivity of the sample proportion assigned to ED-related classes to alternative groupings and posterior weighting.

| Exploratory grouping | Classes grouped as ED-related | Rationale | n | Sample (%) |
|---|---|---|---|---|
| Primary six-class grouping | LC1, LC2, LC4 | Primary profile-based grouping. | 456 | 60.9 |
| Primary six-class grouping, posterior-weighted | LC1, LC2, LC4 | Sum of posterior probabilities instead of most likely class assignment. | 453.8[a] | 60.6 |
| Five-class solution | Three classes (n=219, 71, and 198) | These classes showed mean probabilities of approximately .58 or higher in at least two of the three emotion-related domains. | 488 | 65.2 |
| Excluding LC4 | LC1, LC2 | LC4 showed a low mean probability in the temper domain (.25). | 252 | 33.6 |
| LC2 only | LC2 | The only class with high mean probabilities across all three emotion-related domains. | 184 | 24.6 |
| Including LC6 | LC1, LC2, LC4, LC6 | LC6 had the highest mean probability across the three emotion-related domains among the remaining classes. | 577 | 77.0 |

**Note.** Except for the posterior-weighted row, counts and percentages are based on assignment to the most likely latent class. The posterior-weighted expected count is the sum of individual posterior probabilities for the included classes; its percentage is based on N=749. All groupings are exploratory and should not be interpreted as estimates of the clinical prevalence of emotional dysregulation. ED, emotional dysregulation; LC, latent class.

[a] Expected count based on posterior class probabilities.

### ED and comorbid disorders

**Table S10**. The 14 personality styles from the PSSI by group.

| **Personality styles** (M±SD) | **All patients** | **ADHD** | **ADHD+ED** | **Test/ES** | ***p*** | **Adjusted *p*** |
|---|---|---|---|---|---|---|
| Paranoid | 49.1±12.8 | 46.5±12.5 | 50.8±12.8 | Z=4.51; *r*=.17 | <.001 | <.001 |
| Schizoid | 52.9±11.5 | 52.7±10.8 | 53.0±12.0 | Z=1.03; *r*=.04 | .304 | .328 |
| Schizotypal | 43.7±10.5 | 42.1±10.0 | 44.7±10.6 | Z=3.05; *r*=.12 | .002 | .003 |
| Borderline | 54.7±9.5 | 51.0±9.0 | 57.0±9.1 | Z=8.29; *r*=.32 | <.001 | <.001 |
| Histrionic | 51.1±11.1 | 49.7±10.5 | 52.0±11.4 | Z=2.80; *r*=.11 | .005 | .006 |
| Narcissistic | 51.6±12.3 | 49.8±10.7 | 52.7±13.1 | Z=4.44; *r*=.17 | <.001 | <.001 |
| Avoidant | 55.4±10.6 | 53.0±10.8 | 57.0±10.2 | Z=5.10; *r*=.20 | <.001 | <.001 |
| Dependent | 52.4±10.2 | 50.8±9.9 | 53.5±10.2 | Z=3.53; *r*=.14 | <.001 | <.001 |
| Obsessive-compulsive | 46.9±10.3 | 48.5±9.8 | 45.8±10.5 | Z=−3.31; *r*=.13 | <.001 | .001 |
| Negativistic | 54.0±10.9 | 50.1±10.2 | 56.4±10.6 | Z=7.18; *r*=.28 | <.001 | <.001 |
| Depressive | 57.2±9.7 | 54.7±9.1 | 58.8±9.7 | Z=6.03; *r*=.23 | <.001 | <.001 |
| Self-sacrificing | 52.9±10.4 | 52.0±9.8 | 53.5±10.7 | Z=2.09; *r*=.08 | .037 | .043 |
| Rhapsodic | 49.5±10.8 | 49.1±10.7 | 49.7±10.9 | Z=0.78; *r*=.03 | .436 | .436 |
| Antisocial | 49.8±11.8 | 47.9±10.3 | 51.0±12.5 | Z=3.44; *r*=.13 | <.001 | .001 |

**Note.** ADHD, group assigned to the remaining latent classes; ADHD+ED, exploratory group assigned to classes with elevated probabilities in emotion-related WRAADDS domains; ED, emotional dysregulation; PSSI, Personality Style and Disorder Inventory; M, mean; SD, standard deviation; ES, effect size; r, Mann–Whitney effect size. T scores are reported. Group differences were tested using Mann–Whitney U tests. Adjusted p-values were calculated using the Benjamini–Hochberg procedure within the PSSI test family. PSSI analyses were exploratory. The number of included cases varied by subscale: paranoid, 667; schizoid, 670; schizotypal, 673; borderline, 671; histrionic, 670; narcissistic, 672; avoidant, 671; dependent, 673; obsessive-compulsive, 668; negativistic, 670; depressive, 670; self-sacrificing, 670; rhapsodic, 672; antisocial, 671.

### ED and functional impairment

**Table S11**. Exploratory FLRS subscale scores by group.

| **FLRS subscale** (M±SD) | **All patients** | **ADHD** | **ADHD+ED** | **Test/ES** | ***p*** | **Adjusted *p*** |
|---|---|---|---|---|---|---|
| Task Organization | 14.2±3.6 | 13.3±3.6 | 14.7±3.5 | Z=5.14; *r*=.20 | <.001 | <.001 |
| Task Initiation | 14.0±4.4 | 13.3±4.4 | 14.5±4.3 | Z=3.85; *r*=.15 | <.001 | <.001 |
| Task Completion | 12.8±4.0 | 11.5±4.0 | 13.7±3.8 | Z=6.96; *r*=.27 | <.001 | <.001 |
| Time Management | 12.0±4.1 | 10.6±3.8 | 12.8±4.1 | Z=6.97; *r*=.27 | <.001 | <.001 |
| Planning Skills/Goal Planning | 11.5±4.2 | 10.3±4.0 | 12.2±4.2 | Z=5.63; *r*=.22 | <.001 | <.001 |
| Extracting Information | 12.1±3.8 | 11.1±3.8 | 12.8±3.6 | Z=5.62; *r*=.22 | <.001 | <.001 |
| Working Memory | 13.5±4.2 | 12.8±4.2 | 14.0±4.0 | Z=3.67; *r*=.14 | <.001 | <.001 |
| Social Functioning | 10.1±3.6 | 9.2±3.3 | 10.6±3.6 | Z=5.27; *r*=.20 | <.001 | <.001 |

**Note**. ADHD, group assigned to the remaining latent classes; ADHD+ED, exploratory group assigned to classes with elevated probabilities in emotion-related WRAADDS domains; ED, emotional dysregulation; FLRS, Functional Level Rating Scale; M, mean; SD, standard deviation; ES, effect size; r, Mann–Whitney effect size. FLRS subscale names were translated from German for reporting purposes; each subscale comprises two items assessing functioning in everyday task performance. Group differences were tested using Mann–Whitney U tests. Adjusted p-values were calculated using the Benjamini–Hochberg procedure within the FLRS subscale test family. These subscale analyses were exploratory. The number of included cases varied by subscale: Task Organization, 675; Task Initiation, 674; Task Completion, 675; Time Management, 676; Planning Skills/Goal Planning, 676; Extracting Information, 676; Working Memory, 676; Social Functioning, 671.

### *Sensitivity analysis*

**Table S12**. Five-class sensitivity analyses of group differences.

**A.** Demographic and clinical characteristics by group.

| Variable | ADHD | ADHD+ED | Test/ES | *p* | Adjusted *p* |
|---|---|---|---|---|---|
| N (%) | 261 (34.8) | 488 (65.2) | — | — | — |
| Age in years (M±SD) | 33.7±10.8 | 33.6±9.7 | Z=0.36; r=.01 | .721 | .721 |
| Gender, n (%) | | | V=.067 | .199 | .278 |
| Male | 154 (59.0) | 254 (52.0) | | | |
| Female | 105 (40.2) | 229 (46.9) | | | |
| Diverse | 2 (0.8) | 5 (1.0) | | | |
| ADHD presentation, n (%)[a] | | | V=.213 | <.001 | <.001 |
| Inattention | 32 (13.6) | 17 (3.7) | | | |
| Hyperactive-impulsive | 3 (1.3) | 2 (0.4) | | | |
| Combined | 197 (83.8) | 438 (95.8) | | | |
| Unclassified | 3 (1.3) | 0 (0.0) | | | |
| WRAADDS Scores (M±SD)[b] | | | — | — | — |
| Attention deficit | 7.5±1.6 | 8.8±1.1 | | | |
| Hyperactivity/restlessness | 3.2±1.5 | 4.7±1.2 | | | |
| Temper | 2.5±1.7 | 4.2±1.5 | | | |
| Affective lability | 4.9±1.8 | 6.4±1.5 | | | |
| Emotional overreactivity | 3.0±1.7 | 4.5±1.5 | | | |
| Disorganization | 6.3±1.8 | 8.3±1.4 | | | |
| Impulsivity | 5.4±1.9 | 7.4±1.8 | | | |

**B**. Sociodemographic characteristics by group.

| Variable | ADHD | ADHD+ED | Test/ES | *p* | Adjusted *p* |
|---|---|---|---|---|---|
| Family status, n (%) | | | V=.110 | .166 | .278 |
| Single | 72 (27.6) | 165 (33.8) | | | |
| Partnership/married | 130 (49.8) | 244 (50.0) | | | |
| Divorced | 10 (3.8) | 15 (3.1) | | | |
| Widowed | 0 (0.0) | 3 (0.6) | | | |
| Other/unknown | 49 (18.8) | 61 (12.5) | | | |
| Highest school qualification[c], n (%) | | | V=.103 | .300 | .350 |
| No formal qualification | 4 (1.5) | 10 (2.0) | | | |
| Secondary school certification | 85 (32.6) | 185 (37.9) | | | |
| University entrance qualification | 161 (61.7) | 260 (53.3) | | | |
| Special needs school certification | 0 (0.0) | 5 (1.0) | | | |
| Other/unknown | 11 (4.2) | 28 (5.7) | | | |
| Education, n (%) | | | V=.109 | .167 | .278 |
| No education | 25 (9.6) | 58 (11.9) | | | |
| In vocational training | 11 (4.2) | 14 (2.9) | | | |
| In higher education | 49 (18.8) | 76 (15.6) | | | |
| Completed vocational training | 90 (34.5) | 210 (43.0) | | | |
| Completed university degree | 80 (30.7) | 118 (24.2) | | | |
| Other/unknown | 6 (2.3) | 12 (2.5) | | | |
| Employment status, n (%) | | | V=.120 | .052 | .182 |
| Unemployed | 40 (15.3) | 86 (17.6) | | | |
| Employed | 132 (50.6) | 233 (47.7) | | | |
| Student or in training | 76 (29.1) | 115 (23.6) | | | |
| Retired | 2 (0.8) | 10 (2.0) | | | |
| Other/unknown | 11 (4.2) | 44 (9.0) | | | |

**C**. Psychiatric comorbidities and related clinical measures by group.

| **Variable** | **ADHD** | **ADHD+ED** | **Test/ES** | ***p*** | **Adjusted *p*** |
|---|---|---|---|---|---|
| Presence comorbid disorder, n (%) | 204 (78.2) | 392 (80.3) | OR=1.14 | .506 | .506 |
| Number of Comorbidities (M±SD) | 1.0±0.7 | 1.2±0.9 | Z=3.11; *r*=.11 | .002 | .003 |
| Comorbid disorders[d], n (%) | | | V=.214 | <.001 | <.001 |
| No current comorbid disorder | 74 (28.4) | 147 (30.1) | | | |
| Depressive disorder | 60 (23.0) | 81 (16.6) | | | |
| Adjustment disorder | 59 (22.6) | 64 (13.1) | | | |
| Substance use disorder | 14 (5.4) | 45 (9.2) | | | |
| Anxiety disorder | 21 (8.0) | 35 (7.2) | | | |
| Other comorbid disorders | 9 (3.4) | 9 (1.8) | | | |
| Multiple comorbid disorders | 24 (9.2) | 107 (21.9) | | | |
| Questionnaires (M±SD) | | | | | |
| BDI-II sum score | 14.0±10.0 | 18.0±11.5 | Z=4.42; *r*=.17 | <.001 | <.001 |
| AUDIT sum score | 5.5±5.0 | 5.8±5.5 | Z=0.21; *r*=.01 | .832 | .832 |
| DUDIT sum score | 3.4±5.7 | 4.6±7.2 | Z=1.15; *r*=.05 | .250 | .264 |

**D**. Symptom severity and functional impairment by group.

| **Variable** | **ADHD** | **ADHD+ED** | **Test/ES** | ***p*** | **Adjusted *p*** |
|---|---|---|---|---|---|
| CAARS (M±SD) | | | | | |
| Inattention/cognitive problems | 72.5±11.9 | 79.1±9.8 | Z=7.18; r=.27 | <.001 | <.001 |
| Hyperactivity/restlessness | 65.3±13.2 | 71.4±13.1 | Z=5.72; r=.22 | <.001 | <.001 |
| Impulsivity/emotional lability | 61.2±12.3 | 70.2±13.0 | Z=8.24; r=.32 | <.001 | <.001 |
| Problems with self-concept | 64.7±12.8 | 69.2±12.9 | Z=4.38; r=.17 | <.001 | <.001 |
| ADHD-SR (M±SD) | | | | | |
| Total score | 28.4±8.4 | 34.7±7.5 | Z=9.28; r=.35 | <.001 | <.001 |
| Inattentive score | 15.8±4.6 | 18.8±4.1 | Z=8.38; r=.32 | <.001 | <.001 |
| Hyperactive score | 7.1±3.5 | 8.9±3.3 | Z=6.41; r=.24 | <.001 | <.001 |
| Impulsive score | 5.5±3.1 | 7.0±2.8 | Z=5.90; r=.22 | <.001 | <.001 |
| ASRS-v1.1 (M±SD) | | | | | |
| Total score | 45.4±8.6 | 51.8±7.3 | Z=9.56; r=.36 | <.001 | <.001 |
| Inattentive score | 24.7±4.5 | 27.6±3.9 | Z=8.32; r=.32 | <.001 | <.001 |
| Hyperactive/impulsive score | 20.6±6.0 | 24.2±5.1 | Z=7.81; r=.29 | <.001 | <.001 |
| WURS-K total score (M±SD) | 34.1±13.1 | 39.7±12.9 | Z=5.40; r=.20 | <.001 | <.001 |
| SCL-90-R (M±SD) | | | | | |
| Global severity index | 60.3±8.4 | 64.1±7.6 | Z=5.82; r=.23 | <.001 | <.001 |
| Positive symptom total | 57.9±7.8 | 61.2±7.1 | Z=5.23; r=.20 | <.001 | <.001 |
| Positive symptom distress index | 61.5±7.7 | 64.1±7.6 | Z=4.28; r=.17 | <.001 | <.001 |
| General functional level (M±SD) | 5.2±2.0 | 5.8±2.2 | Z=3.86; r=.15 | <.001 | <.001 |
| Documented lifetime suicide attempts, n | 7 | 18 | — | — | — |

**Note.** ADHD+ED, exploratory group assigned to the three classes of the five-class solution with elevated probabilities in emotion-related WRAADDS domains (n=219, 71, and 198; Supplementary Table S9); ADHD, group assigned to the remaining classes. These labels do not denote clinical diagnoses. Instrument abbreviations as in Supplementary Table S1; WRAADDS, Wender-Reimherr Adult Attention Deficit Disorder Scale; M, mean; SD, standard deviation; ES, effect size; OR, odds ratio; r, Mann–Whitney effect size; V, Cramér's V. Tests: Mann–Whitney U (continuous/ordinal), Fisher's exact (2 × 2), exact Fisher–Freeman–Halton (larger tables). Adjusted p, Benjamini–Hochberg-adjusted within the test families described in the Methods. Available-case sample sizes and category definitions as in Tables 2–5. Suicide attempts were not tested. Compared with the six-class grouping, only the DUDIT total score was no longer significant after correction.

[a] Available for N=692 (ADHD=235; ADHD+ED=457); three participants did not meet the questionnaire criteria for a presentation.

[b] Descriptive only; no inferential tests were conducted.

[c] German education system; "secondary school certification" includes lower and middle school-leaving certificates.

[d] Categories with a frequency below 1.5% were grouped as other comorbid disorders. Presence of a comorbid disorder includes 68 participants with recurrent depressive disorder in remission (ICD-10 F33.4), classified as "no current comorbid disorder" in the grouped categories.

**Table S13.** Missing self-report data by instrument and group.

| Instrument | Complete (n) | Incomplete (n) | Missing (%) | | OR | *p* | Adjusted *p* |
|---|---|---|---|---|---|---|---|
| | | | ADHD | ADHD+ED | | | |
| CAARS (IA/HY/SK) | 687 | 62 | 8.9 | 7.9 | 0.88 | .684 | .889 |
| CAARS (IEL) | 683 | 66 | 9.9 | 8.1 | 0.80 | .429 | .726 |
| ADHD-SR | 692 | 57 | 9.6 | 6.4 | 0.64 | .121 | .339 |
| ASRS-v1.1 | 699 | 50 | 7.8 | 5.9 | 0.74 | .299 | .589 |
| WURS-K | 699 | 50 | 8.5 | 5.5 | 0.62 | .133 | .358 |
| SCL-90-R | 667 | 82 | 10.9 | 11.0 | 1.00 | 1.000 | 1.000 |
| BDI-II | 717 | 32 | 4.1 | 4.4 | 1.07 | 1.000 | 1.000 |
| AUDIT | 626 | 123 | 12.6 | 18.9 | 1.61 | .026 | .124 |
| DUDIT | 622 | 127 | 13.0 | 19.5 | 1.63 | .022 | .110 |
| FLRS | 661 | 88 | 11.3 | 12.1 | 1.08 | .816 | .954 |
| PSSI (all 14 scales) | 645 | 104 | 15.7 | 12.7 | 0.78 | .279 | .567 |

**Note.** ADHD+ED, exploratory group assigned to latent classes with elevated probabilities in emotion-related WRAADDS domains (n=456); ADHD, group assigned to the remaining classes (n=293). Instrument abbreviations as in Supplementary Table S1; CAARS IA/HY/SK, inattention/cognitive problems, hyperactivity/restlessness, and problems with self-concept (scales with identical missing-data patterns, combined); CAARS IEL, impulsivity/emotional lability; WRAADDS, Wender-Reimherr Adult Attention Deficit Disorder Scale. Missing (%), percentage of participants with missing data in each group; OR, odds ratio for missing data in the ADHD+ED versus the ADHD group; p, Fisher's exact test; Adjusted p, Benjamini–Hochberg-adjusted across all missing-data tests and completer–non-completer comparisons; r, Mann–Whitney effect size. For the PSSI, completers had all 14 scale scores (n per scale: 667–673). Completers and non-completers were also compared on age, gender, year of assessment, WRAADDS domain scores, and number of comorbid diagnoses. Year-of-assessment differences were observed for most instruments ($r$=.11–.40); gender distributions differed for the AUDIT, DUDIT, and PSSI (Cramér's $V$=.12–.18). WRAADDS domain differences were small ($r \leq .13$), and no differences in age or number of comorbid diagnoses remained significant after correction.

**Table S14.** Core-adjusted group comparisons and dimensional associations.

**A.** Adjusted group comparisons.

| **Variable** | **n** | **Unadjusted model** Est. (95% CI); std. | **Adj.** ***p*** | **Core-adjusted model** Est. (95% CI); std. | **Adj.** ***p*** | **Rank ANCOVA Adj.** ***p*** |
|---|---|---|---|---|---|---|
| CAARS | | | | | | |
| Inattention/cognitive problems | 687 | 7.10 (5.41, 8.78); 0.65 | <.001 | 2.69 (0.46, 4.93); 0.25 | .043 | .010 |
| Hyperactivity/restlessness | 687 | 6.59 (4.58, 8.59); 0.49 | <.001 | −2.99 (−5.63, −0.35); −0.22 | .056 | .066 |
| Impulsivity/emotional lability | 683 | 10.45 (8.53, 12.36); 0.78 | <.001 | 4.36 (1.71, 7.00); 0.32 | .005 | .007 |
| Problems with self-concept | 687 | 5.42 (3.45, 7.40); 0.42 | <.001 | 4.81 (2.08, 7.53); 0.37 | .003 | <.001 |
| ADHD-SR | | | | | | |
| Total score | 692 | 6.79 (5.57, 8.00); 0.81 | <.001 | −0.11 (−1.59, 1.37); −0.01 | .934 | .780 |
| Inattentive score | 692 | 3.28 (2.62, 3.94); 0.73 | <.001 | 0.63 (−0.28, 1.54); 0.14 | .300 | .049 |
| Hyperactive score | 692 | 1.86 (1.35, 2.38); 0.54 | <.001 | −0.60 (−1.28, 0.08); −0.17 | .162 | .157 |
| Impulsive score | 692 | 1.64 (1.19, 2.09); 0.56 | <.001 | −0.14 (−0.74, 0.45); −0.05 | .724 | .544 |
| ASRS-v1.1 | | | | | | |
| Total score | 699 | 6.87 (5.66, 8.08); 0.83 | <.001 | −0.41 (−1.83, 1.01); −0.05 | .724 | .894 |
| Inattentive score | 699 | 3.03 (2.38, 3.67); 0.70 | <.001 | 0.19 (−0.62, 1.00); 0.04 | .724 | .316 |
| Hyperactive/impulsive score | 699 | 3.84 (3.00, 4.69); 0.67 | <.001 | −0.60 (−1.70, 0.51); −0.10 | .426 | .544 |
| WURS-K total | 699 | 6.58 (4.62, 8.54); 0.50 | <.001 | 1.42 (−1.22, 4.06); 0.11 | .426 | .733 |
| SCL-90-R | | | | | | |
| Global severity index | 667 | 4.43 (3.20, 5.66); 0.55 | <.001 | 3.12 (1.46, 4.79); 0.39 | .002 | <.001 |
| Positive symptom total | 666 | 3.70 (2.55, 4.85); 0.49 | <.001 | 2.41 (0.88, 3.93); 0.32 | .006 | .001 |
| Positive symptom distress index | 667 | 3.50 (2.32, 4.68); 0.45 | <.001 | 2.52 (0.86, 4.18); 0.33 | .008 | .002 |
| General functional level | 661 | 0.80 (0.48, 1.12); 0.38 | <.001 | 0.78 (0.34, 1.23); 0.37 | .003 | .001 |
| Questionnaires | | | | | | |
| BDI-II sum score | 717 | 4.70 (3.11, 6.29); 0.42 | <.001 | 4.59 (2.52, 6.65); 0.41 | <.001 | <.001 |
| AUDIT sum score | 626 | 0.45 (−0.38, 1.28); 0.08 | .288 | −0.50 (−1.59, 0.59); −0.09 | .503 | .411 |
| DUDIT sum score | 622 | 1.48 (0.46, 2.51); 0.22 | .005 | 0.00 (−1.40, 1.40); 0.00 | .999 | .551 |
| Presence comorbid disorder | 749 | OR 1.11 (0.78, 1.60) | .559 | OR 1.04 (0.63, 1.72) | .878 | — |
| Number of Comorbidities | 749 | 0.22 (0.11, 0.33); 0.27 | <.001 | 0.15 (−0.02, 0.32); 0.19 | .149 | .218 |

**B.** Dimensional associations.

| Variable | n | Temper | Affective lability | Emotional overreactivity |
|---|---|---|---|---|
| CAARS | | | | |
| Inattention/cognitive problems | 687 | −.14 (<.001) | .03 (.480) | .12 (.003) |
| Hyperactivity/restlessness | 687 | .05 (.202) | .08 (.060) | −.11 (.007) |
| Impulsivity/emotional lability | 683 | .38 (<.001) | .20 (<.001) | .11 (.006) |
| Problems with self-concept | 687 | .02 (.605) | .13 (.002) | .23 (<.001) |
| ADHD-SR | | | | |
| Total score | 692 | .10 (.017) | .06 (.133) | .08 (.071) |
| Inattentive score | 692 | −.07 (.100) | .01 (.769) | .22 (<.001) |
| Hyperactive score | 692 | .16 (<.001) | .07 (.097) | −.06 (.150) |
| Impulsive score | 692 | .18 (<.001) | .06 (.169) | −.03 (.452) |
| ASRS-v1.1 | | | | |
| Total score | 699 | .02 (.718) | .08 (.051) | .02 (.652) |
| Inattentive score | 699 | −.14 (<.001) | .02 (.605) | .13 (.002) |
| Hyperactive/impulsive score | 699 | .14 (<.001) | .09 (.034) | −.06 (.138) |
| WURS-K total | 699 | .23 (<.001) | .06 (.132) | .01 (.822) |
| SCL-90-R | | | | |
| Global severity index | 667 | .16 (<.001) | .18 (<.001) | .20 (<.001) |
| Positive symptom total | 666 | .11 (.007) | .18 (<.001) | .20 (<.001) |
| Positive symptom distress index | 667 | .12 (.005) | .13 (.002) | .11 (.011) |
| General functional level | 661 | .00 (.925) | .12 (.005) | .22 (<.001) |
| Questionnaires | | | | |
| BDI-II sum score | 717 | .11 (.009) | .19 (<.001) | .19 (<.001) |
| AUDIT sum score | 626 | .01 (.844) | .12 (.007) | −.12 (.006) |
| DUDIT sum score | 622 | .01 (.911) | .12 (.005) | −.08 (.067) |
| Presence comorbid disorder | 749 | .00 (.977) | .08 (.116) | .05 (.216) |
| Number of Comorbidities | 749 | .04 (.282) | .09 (.095) | .07 (.120) |

**Note.** ADHD+ED (n = 456) and ADHD (n = 293) denote the exploratory six-class grouping and are not clinical diagnoses. The core symptom score is the mean of the z-standardized WRAADDS domain scores for attention difficulties, hyperactivity/restlessness, disorganization, and impulsivity; emotion-related domains were excluded. In Panel A, estimates are for ADHD+ED minus ADHD. Linear-model estimates with HC3 standard errors are shown in raw units with 95% CIs; the value after the semicolon is the standardized difference in outcome SD units. For the presence of a comorbid disorder, estimates are odds ratios with Wald 95% CIs from logistic regression. Rank-transformed analysis of covariance (ANCOVA) used the ranked outcome and ranked core symptom score with HC3 standard errors; it was not applicable to the binary outcome. In Panel B, entries are partial Spearman correlations (ρ) controlling for the core symptom score, followed in parentheses by adjusted p. Adjusted p-values were Benjamini–Hochberg-adjusted within the clinical and comorbidity families, separately for each analysis; in Panel B, adjustment was applied jointly across all outcomes and the three emotion-related domains within each family. Analyses included participants with nonmissing values for the variables required by each model. Instrument abbreviations are defined in Supplementary Table S1; CI, confidence interval; HC3, heteroscedasticity-consistent covariance estimator; OR, odds ratio; SD, standard deviation; WRAADDS, Wender-Reimherr Adult Attention Deficit Disorder Scale.

**Table S15.** Internal consistency (Cronbach's α) in the present sample.

| Variable | Items | *n* | α |
|---|---|---|---|
| CAARS | | | |
| Inattention/cognitive problems | 12 | 672 | .83 |
| Hyperactivity/restlessness | 12 | 644 | .87 |
| Impulsivity/emotional lability | 12 | 669 | .88 |
| Problems with self-concept | 6 | 679 | .83 |
| ADHD-SR | | | |
| Total score | 18 | 669 | .82 |
| Inattentive score | 9 | 676 | .73 |
| Hyperactive score | 5 | 688 | .75 |
| Impulsive score | 4 | 688 | .76 |
| ASRS-v1.1 | | | |
| Total score | 18 | 672 | .80 |
| Inattentive score | 9 | 693 | .70 |
| Hyperactive/impulsive score | 9 | 681 | .78 |
| WURS-K | 21 | 643 | .87 |
| SCL-90-R, all 90 items | 90 | 555 | .97 |
| Questionnaires | | | |
| BDI-II sum score | 21 | 693 | .91 |
| AUDIT sum score | 10 | 553 | .83 |
| DUDIT sum score | 11 | 595 | .91 |
| FLRS[a] | | | |
| Task organization | 2 | 667 | .57 |
| Task initiation | 2 | 671 | .78 |
| Task completion | 2 | 674 | .55 |
| Time management | 2 | 674 | .50 |
| Planning skills/goal planning | 2 | 666 | .68 |
| Extracting information | 2 | 674 | .48 |
| Working memory | 2 | 673 | .72 |
| Social functioning | 2 | 662 | .53 |
| WRAADDS | | | |
| Attention deficit | 5 | 749 | .44 |
| Hyperactivity/restlessness | 3 | 749 | .58 |
| Temper | 3 | 749 | .75 |
| Affective lability | 4 | 749 | .58 |
| Emotional overreactivity | 3 | 749 | .71 |
| Disorganization | 5 | 749 | .55 |
| Impulsivity | 5 | 749 | .52 |

Note. α, Cronbach's alpha based on complete item responses for each scale, without imputation; n may therefore be smaller than the number of participants with an available scale score. For the SCL-90-R, α was calculated across all 90 items used to derive the GSI, PST, and PSDI; it was not calculated separately for the symptom subscales. [a] Each FLRS subscale comprises two items; estimates based on two items should be interpreted with caution. α could not be estimated for the PSSI because item-level data were unavailable, nor for the general functional-level rating because it is a single item. WRAADDS interrater reliability could not be assessed. Instrument abbreviations are defined in Supplementary Table S1; GSI, Global Severity Index; PST, Positive Symptom Total; PSDI, Positive Symptom Distress Index; WRAADDS, Wender-Reimherr Adult Attention Deficit Disorder Scale.

### *Supplementary Figures*

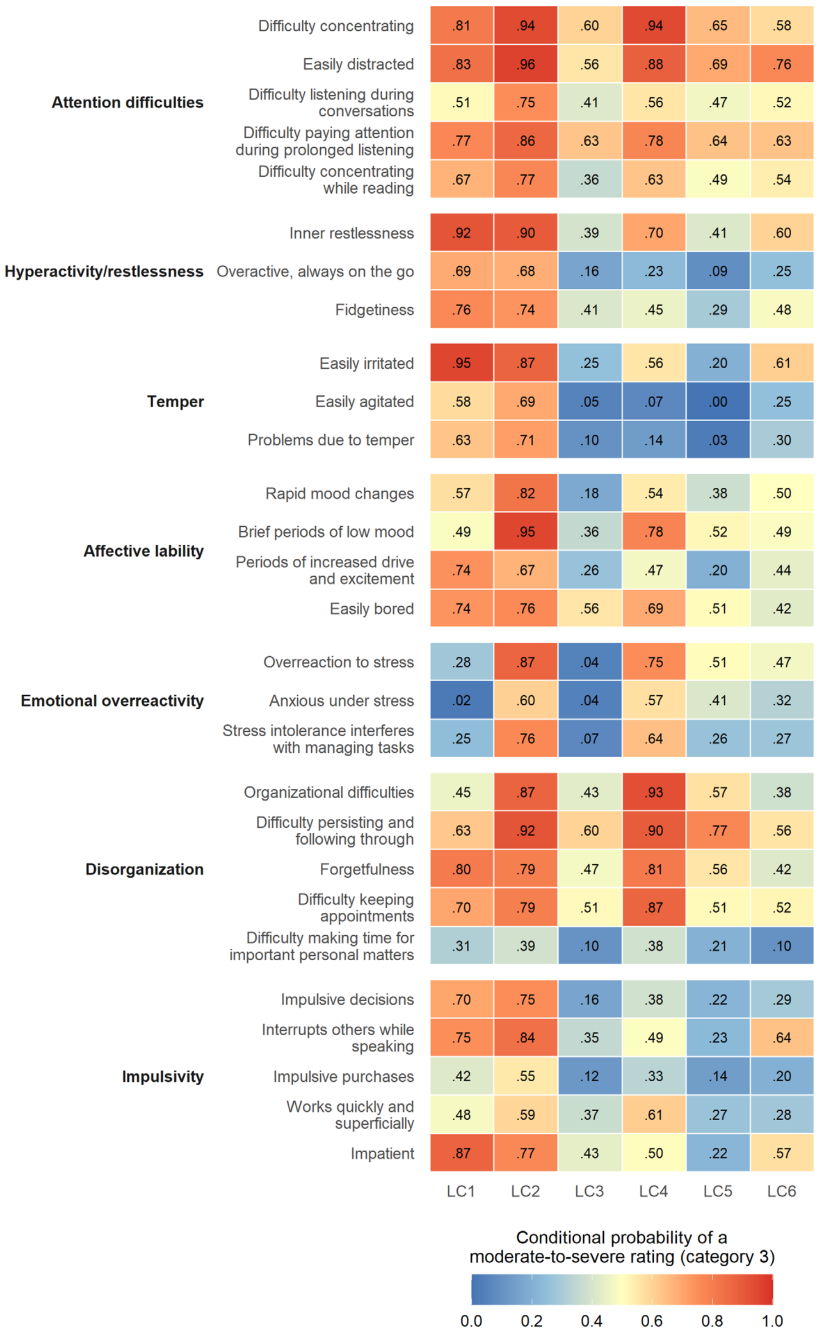


**Figure S1.** Heatmap of class-conditional probabilities of a moderate-to-severe rating (category 3) for the 28 WRAADDS items across six latent classes (LC1–LC6). Rows show items grouped by symptom domain, labeled on the left; columns represent latent classes. Cell colors and overlaid values indicate the class-conditional probability of a category 3 rating (0=blue, 1=red). The exploratory ED-related interpretation was based on elevated probabilities in the temper, affective lability, and emotional overreactivity domains; corresponding domain means are reported in Supplementary Table S8. Descriptive class labels: LC1, HI+ED; LC2, IA/HI+ED; LC3, PR; LC4, IA+ED; LC5, IA; LC6, HI. ED, emotional dysregulation; HI, hyperactivity/impulsivity; IA, inattention; PR, partially remitting symptoms; WRAADDS, Wender-Reimherr Adult Attention Deficit Disorder Scale.
